# A framework for chronic care quality: results of a scoping review and Delphi survey

**DOI:** 10.1101/2024.08.21.24312364

**Authors:** Grace Marie V. Ku, Willem Van De Put, Deogratias Katsuva, Mohamad Ali Ag Ahmed, Megumi Rosenberg, Bruno Meessen

## Abstract

Frameworks conceptualising quality of care abound and vary; some concentrate on specific aspects (e.g., safety, access, effectiveness), others all-encompassing. However, to our knowledge, tailoring to systematically arrive at a comprehensive care for chronic conditions quality (CCCQ) framework has never been done. We conducted scoping review and Delphi survey to produce a CCCQ framework, comprehensively delineating aims, determinants and measurable attributes.

With the assumption that specific groups (people with chronic conditions, care providers, financiers, policy-makers, etc) view quality of care differently, we analysed 48 scientific and 26 grey literature deductively and inductively using the Institute of Medicine’s quality of care framework as the foundation. We produced a zero-version of the quality of chronic care framework, detailing aims, healthcare system determinants, and measurement mechanisms. This was presented in a Delphi survey to 49 experts with diverse chronic care expertise/experience around the world. Consensus was obtained after the first round, with the panel providing suggestions and justifications to expand the agreed-upon components. Through this exercise, a comprehensive CCCQ framework encompassing the journey through healthcare of people with chronic conditions was developed. The framework specifies seven CCCQ ‘aims’ and identifies health system determinants which can be acted upon with ‘organising principles’ and measured through chronic care quality ‘attributes’ related to structures, processes and outcomes. Tailoring quality of care based on the nature of the diseases/conditions and considering different views can be done to ensure a comprehensive offer of healthcare services, and towards better outcomes that are acceptable to both the health system and PwCCs.

## BACKGROUND

The underpinnings of care quality emerged in the 19^th^ century, ranging from underscoring the importance of handwashing to prevent infection, to correlating poor living conditions with increased mortality and the establishment of hospital standards to assess healthcare outcomes [1]. In 1966, Donabedian introduced a framework for healthcare quality measurement laying the groundwork for (modern-day) healthcare quality [2]. Near the end of the 20^th^ century, the Institute of Medicine (IOM) called for designing safer health systems, thereby improving quality of care [3]. The early 21^st^ century brought forth a framework for quality of care [4] and a revision [5] by the IOM, identifying six aims of quality in healthcare. It also saw the emergence of care quality frameworks by various (international) agencies, including the World Health Organization (WHO) [6,7].

However, the aforementioned frameworks apply to quality in healthcare in general. Establishing criteria and a quality framework specific for chronic care can be “messy”. Due to chronicity – with most conditions lasting throughout the lifetime of the person – priorities and attention to increase the likelihood of “desired effects” (usually favourable health outcomes) would be different, as compared to acute diseases. Beyond biomedical needs, it is crucial to support the psychosocial aspects of people with chronic conditions (PwCC) for them *to adapt and self-manage in the face of social, physical, and emotional challenges* [8]. The reality of multimorbidity has to be acknowledged. Furthermore, it should be recognised that the main drivers of chronic conditions, including most social, structural, commercial determinants, are beyond the health system. Additionally, there are various interests of different stakeholders and actors (e.g. healthcare providers, policy decision-makers, financiers, regulators, other sectors), which may be congruent or disparate. Diverse contexts, for instance, low-resource settings with competing priorities and facing double/triple burden of disease, would also be influential.

Looking into what factors matter for good-quality healthcare for chronic conditions therefore requires taking different perspectives of various stakeholders and data sources and making use of different lenses (Figure 1).

**Figure 1.**
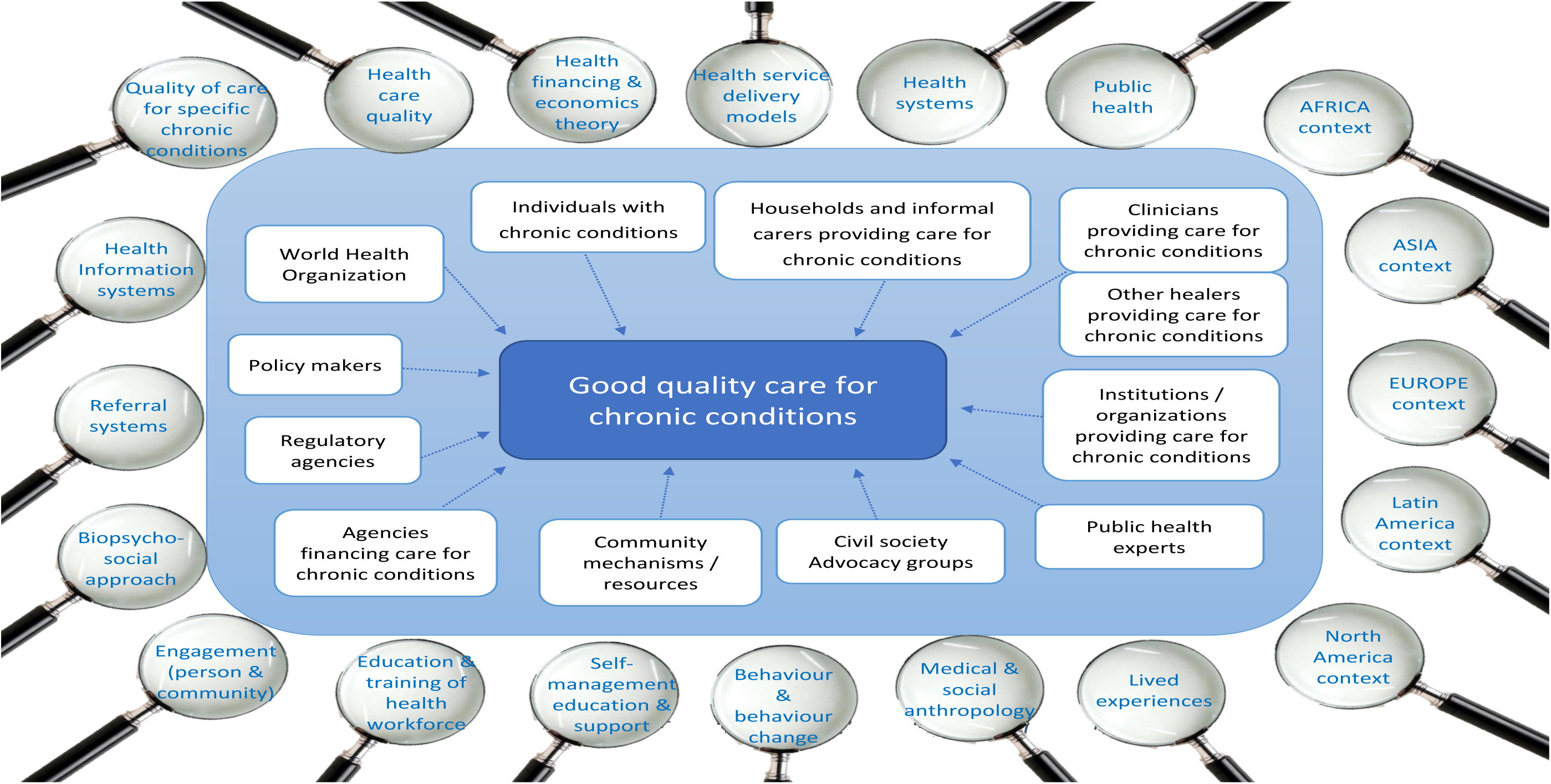
The multi-lens methodological framework

With the above considerations in mind, we conducted this study to produce a framework that comprehensively delineates aims, determinants and measurable attributes of quality of care specifically for chronic conditions.

### Conceptual Issues and Definitions

Instead of ‘reinventing the wheel’, we adopted the latest version of the quality-of-care framework put forward by the Institute of Medicine (5), noting that this is generic and needs to be contextualized.

We defined aims, determinants and attributes as follows:

1. ‘Aim’ – any broad category of importance with intrinsic value, as a desired final outcome that is achieved to denote that care is of good quality. Since the IOM’s framework for quality-of-care reports [4,5], the consensus converges around a list of six aims: effectiveness; efficiency; safety; equity; accessibility, timeliness, affordability; and person-centredness. While we are aware that different documents/reports have extended or reorganized this list, we took the IOM quality aims as our starting point.
2. *‘Determinant’* – any actionable factor which has direct effect on achieving any dimension of quality of care. These can be considered as the ‘elements’ presented in models for chronic care. The determinants also correspond to the “health system determinants” as described by WHO [8] to support a health system in delivering healthcare. Determinants may extend to systemic challenges health system-wise, including arrangements within a system or a model of health service delivery and the conditions/limitations/opportunities to be found in family, community resources, the environment, and the community itself.
3. *Attribute* – any variable of importance that measures achievement of specific quality aim(s) or fulfilment of (some of) its determinants. It can relate to a specific chronic condition or in general. This can be considered as the overall measurable characteristics of the different aims and/or determinants of good quality healthcare, as well as actions on determinants (along organising principles) to achieve the quality aims, for which direct indicators and criteria can be formulated (usually based on context).

## METHODOLOGY

This paper draws specific results from a larger programme of work commissioned by WHO. The request was to produce a comprehensive conceptualization of “quality health services for chronic conditions” that can be used by actors considering interventions to improve health services for chronic conditions, in this case, purchasing arrangements as an instrument for improvement, with a particular attention to policy needs of low- and middle-income countries (LMICs). Here, we concentrate on the components relative to the framework, specifically the determinants, actions and some of their organising principles, and measurable attributes. The chronic care quality aims have already been presented in an earlier paper [9].

We reviewed relevant literature and convened international stakeholders for chronic care and quality in a Delphi survey.

### Scoping review

We conducted a scoping review following the PRISMA extension guidelines [10] to systematically identify available information on quality of care for chronic conditions, identifying key concepts. We selected works that have acknowledged and unpacked the plurality of quality in chronic care, and which proposed/made use of frameworks or looked into two or more IOM aims of care quality and studied or demonstrated implementation. The scoping review protocol is available from https://www.itg.be/en/research/research-themes/quality-of-care-for-chronic-conditions.

### Scientific publications

On 2 February 2022, search for scientific publications was conducted in the PubMed and Science Direct data bases using specific search terms: *‘chronic condition’/’chronic illness’/’chronic disease’; ‘quality of healthcare’; ‘innovative care for chronic conditions’; ‘chronic care model’; ‘quality criteria’; ‘quality indicators’*; specific chronic conditions considered among top drivers of chronic disease burden [11] (*‘ischaemic heart disease’, ‘hypertension’ and ‘stroke’; ‘diabetes mellitus’; ‘chronic kidney disease’; ‘lung cancer’; ‘HIV/AIDS’; ‘chronic obstructive pulmonary disease’ and ‘bronchial asthma’*) and additional conditions as suggested by the WHO team *(‘chronic musculoskeletal conditions’; ‘chronic skin disease’*); and criteria: *written in English or French; publication years 2002-2021; among humans*.

### Other literature and documents

Search for grey/other literature (policies, circulars, publications not available from scientific search engines) were conducted using the same keywords but including general quality of care documents and with broader year limitations (1999-2022) in the Google search engine. Additionally, contacts from the WHO, healthcare regulatory agencies, organizations with chronic disease programs/projects, and various Ministries of Health and/or connected agencies were requested to share any documents they have produced as related to quality of care, specifically for chronic conditions.

### Literature sifting

Scientific publications were sifted through Rayyan (www.rayyan.ai). This was done systematically by minimum two members of the research team with any disagreements resolved amongst the two, as needed, through a third researcher. Retrieved scientific publications were initially screened through the titles. Abstracts (if available) of the chosen documents were individually reviewed. Full articles were scrutinized and selected; only documents that are relevant to this study were included in the final selection. We also looked into the bibliographic references of the included articles to check for additional literature; however, none of the snowballed papers were included in the final list. (Figure 2)

**Figure 2.**
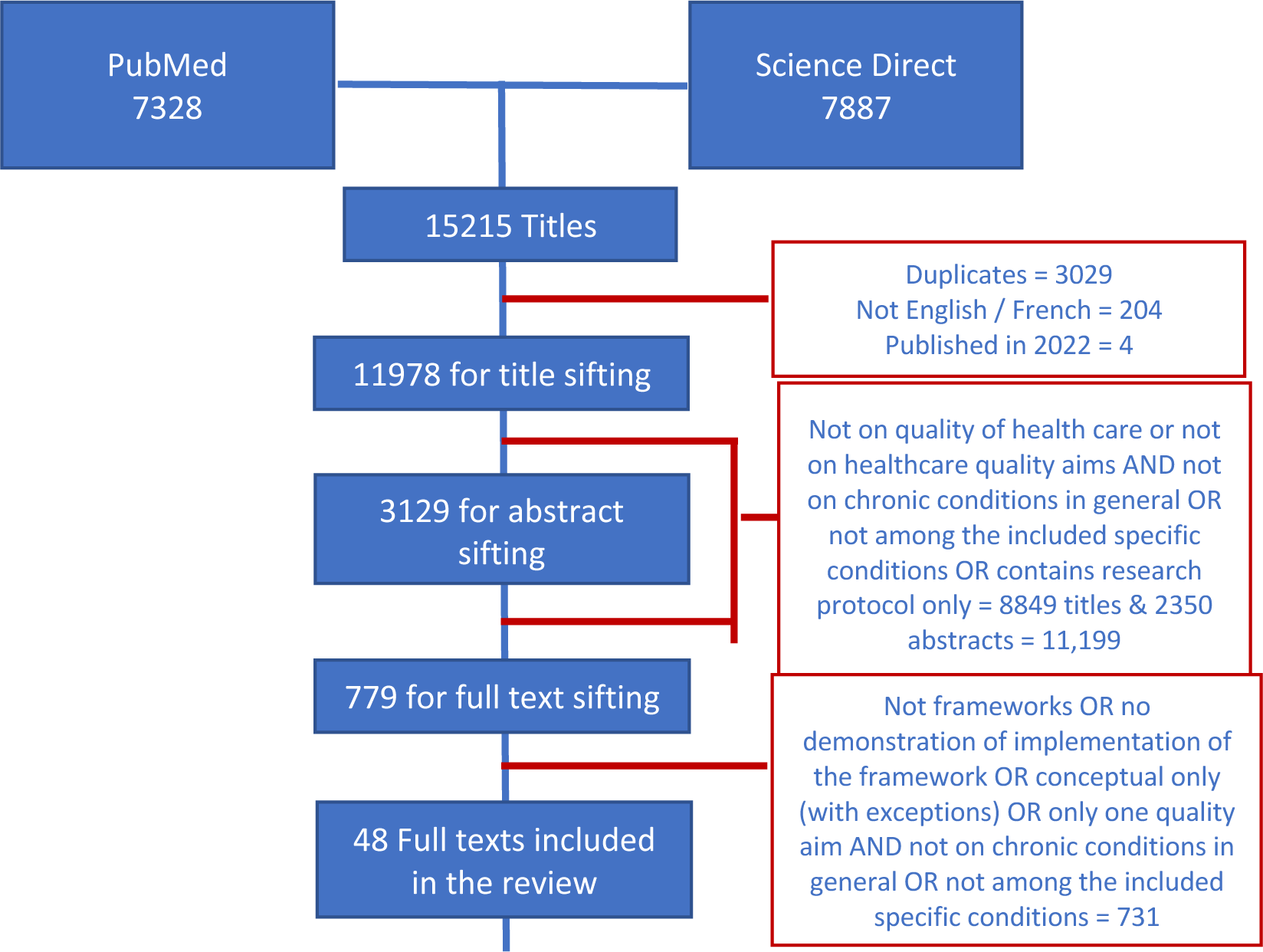
Sifting of retrieved scientific literature

Grey literature and other documents were purposively collected.

### Data extraction and framework building

Data retrieval was systematically initiated by at least one of the members of the research team and verified by a different member. We critically analysed the literature and made use of deductive and inductive approaches to identify quality aims, determinants and attributes. We used our definitions for deductive and inductive analysis. We utilised the IOM quality framework to deductively extract data on aims. Analysis was done iteratively, going back to the literature as we identified additional concepts and critically analysing to expound on the meanings. We intended to group determinants following the WHO health systems building blocks [8]. However, as we also considered resources beyond the health system, we reclassified the groups to: (1) leadership and governance; (2) financing; (3) resources (including health workforce, health information, medical products, vaccines, and technologies, PwCC, their families, the community and other sectors); and (4) service delivery. We then inductively identified specific determinants, any specific actions on these determinants, and corresponding principles organising said actions. We also determined measurable attributes relative to each of the chronic care quality aims. Additionally, during data extraction, we noted that specific papers would concentrate on a particular stage in the ‘journey’ of a person in the natural history of a chronic condition (e.g. addressing risks, rehabilitation, etc). We thus went back to our selection to consciously extract additional information on particular stages of the PwCC journey. We then brought forward said concepts, as related to the aims, determinants, and attributes of good quality chronic care, and the principles that organise actions on the determinants, to build the zero version of the chronic care quality (CCQ) framework. To note, the lived experiences of the research team (as health care provider, PwCC, health economist, anthropologist, public health expert; from LIC, MIC, HIC) influenced the critical analysis of the data.

### Delphi survey

We prepared a list of “mid”- to “advanced” level chronic care experts, gleaned through our own professional networks, references from colleagues or other known experts, relevant publications in peer-reviewed journals or the websites of relevant organizations, and from recommendations by different organisations (e.g. NCD Alliance, Global Alliance on Chronic Diseases), and supplemented by the WHO Team. A concern was to secure, to the extent possible, representation across genders, types of expertise, and settings of activities/experience (with focus on low- and middle-over high-income settings), covering the six WHO regions. We conducted two rounds of the Delphi survey via an online application, Mesydel (https://mesydel.com/en). The first step was to arrive at an agreement over our scoping review findings that build towards the chronic care quality framework, and to propose financing mechanisms to improve quality of chronic care. Based on first round results, the second round was conducted to fine-tune purchasing arrangements. For this paper, we concentrated on findings contributing to the chronic care quality framework. Findings related to financing mechanisms to improve quality of chronic care will be presented in a separate paper.

We presented scoping review findings and version zero of the chronic care quality framework to our Delphi respondents. There was consensus on the identified chronic care quality aims, the groups of determinants, and our proposed actions in the first round. The respondents gave rich suggestions on how each of the chronic care quality aims could be achieved, providing specific determinants and actions based on their own settings, to complement the chronic care quality framework. We synthesised and critically analysed the responses, reflecting on our scoping review findings and contrasting and comparing all information collected.

## RESULTS

A total of 15,215 scientific articles were retrieved and 48 [12–59] were retained for the review (Figure 1). Eighteen of these are specific for certain chronic conditions (diabetes=5, cardiovascular diseases including hypertension and stroke=5, HIV/AIDS=2; chronic obstructive pulmonary disease=2, chronic kidney disease=2, osteoarthritis=1, and cancer=1) while some target specific groups (older people=5, children=1, female=1, informal caregiver=1). Forty-six (46) propose and implement or demonstrate implementations of various models of quality of care, mostly in high income countries (n=31), five in LMICs (South Africa=3, Haiti=1, not specified=1), and the rest (n=10) said to be global/international. Majority (n=46) fit and consolidate the IOM definition of quality and two or more of the IOM care quality aims. A couple[41–42] consider Donabedian’s [2] elements of quality in healthcare.

We retrieved 26 grey literature/documents from the IOM (n=3)[3–5]; WHO (n=10)[7,60–68] the EU Joint Action on Chronic Diseases and Healthy Ageing Across the Life Cycle (n=4)[69–72]; the United States of America (USA) Agency for Health Care Research & Quality (n=2)[73–74]; and the rest coming from different agencies: two from the USA [75–76], and one document each from Australia [77], Canada [78] Ireland [79], Belgium [80],and the Philippines [81].

More detailed information extracted from the scientific and grey literature can be found in the supplementary files, available from https://www.itg.be/en/research/research-themes/quality-of-care-for-chronic-conditions.

### The natural history of (most) chronic conditions and the PwCC journey

Our scoping review findings [5,13–14,29,33,37,52,55,59,64,68] indicate a path of a person who is at risk of and will eventually develop (and die from) a chronic condition. In the natural history of most chronic conditions, exposure of a person throughout the whole life-course (from foetal life to adulthood) to certain social, structural and commercial determinants of health and diverse risk factors predisposes a person to develop chronic conditions. Left unaddressed, continued exposure to risks and determinants will likely cause maldevelopment of and/or damage to various organ systems including the immune system and, eventually, lead to the development of chronic conditions. Using this person’s path through the ‘natural history’ of (most) chronic conditions, we mapped out the journey through healthcare of a person who is at risk of and will eventually develop (and die from) a chronic condition, and expectations from the healthcare system. We propose the stages in the journey and touchpoints in healthcare as follows (see Figure 3):

1. general population, for risk prevention
2. at (high) risk population, for risk control
3. PwCC, of which there would be different touchpoints

a. diagnosis and prompt treatment,
b. follow-up

i. for the condition

(1) regular follow-up
(2) during exacerbations/moments of crisis
ii. for co-morbid conditions
iii. for complications
iv. for other health problems
c. rehabilitation
d. palliative and end-of-life care
e. considerations for the primary informal caregivers of PwCC

**Figure 3.**
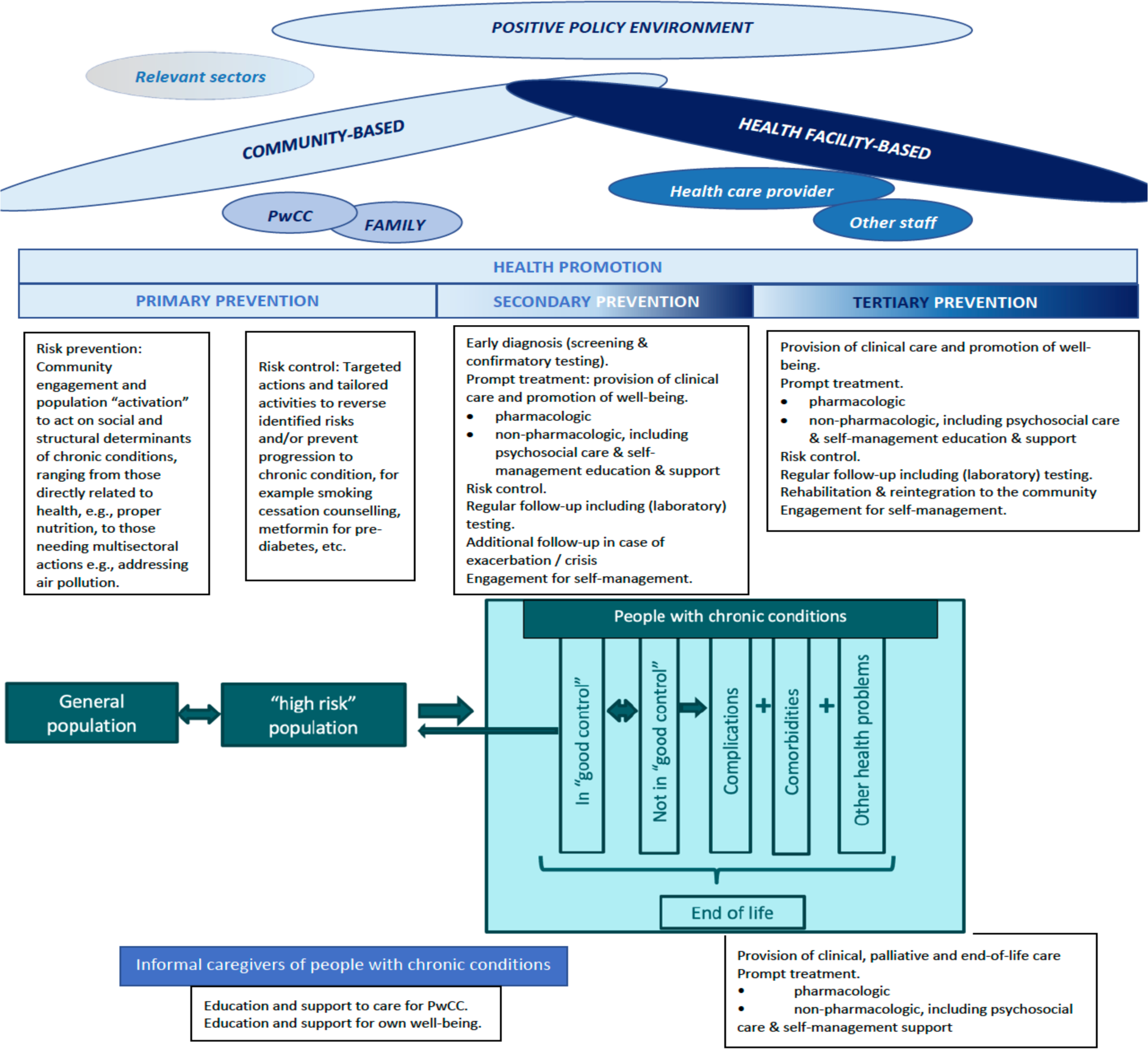
*Journey through the healthcare delivery system of a person with chronic condition (PwCC Journey)* Explanatory notes: **Community-based** services include, among others, general health promotion activities; awareness-raising; risk-screening and control; early diagnosis (screening for disease); self-management support; linking with community resources; community reintegration and community-based rehabilitation. **Facility-based** services include, among others, early diagnosis (confirmatory); prompt and regular clinical case management; self-management education; targeted risk control; for complications and comorbidities/multimorbidities - early recognition (including screening), prompt and regular treatment and referral (as needed). ‘**Good control**’ refers to achievement *of good clinical endpoints* of the chronic condition; logically, ‘**not in good control**’ is the opposite. In a lifetime of a PwCC, it is expected that there will be moments of ‘good’ control and moments of ‘poor’ (not in good) control. The frequency of either, and the severity of ‘poor control’ depends on a number of factors, the quality of chronic care of which plays a vital role. **Comorbidity** is the occurrence of multiple (acute and/or chronic medical conditions which *may or may not be related with the chronic condition* of the PwCC, and which contribute to their total burden of (physiologic as well as psychosocial) dysfunction. While these may either be acute or other chronic conditions, comorbidities are usually used for additional chronic conditions. The presence of comorbidities complexifies healthcare as there may be agonistic and antagonistic interactions in case management. ‘**Complications**’ are conditions that arise from (worsening of) the chronic condition(s) including those that arise during the course of treatment. **Other health problems** are health issues which are unrelated to the chronic condition but are recognised as such because of effects of the chronic condition on the health and well-being of the person. These may also be classified as comorbid conditions, but we use this term to refer to other health problems not necessarily with established diagnosis and may include the need for preventive services, e.g., immunizations, prophylaxis treatments, as warranted by the chronic condition.

The depicted journey and corresponding touchpoints with the healthcare system should not be viewed in a linear, streamlined fashion. People may skip stages, ‘leave’ without completing the whole journey (where death may be from other causes, e.g. a fatal accident), or may regress to a preceding stage. PwCC may experience more than one of the depicted stages (e.g. not in good control + complications + other health problems). Additionally, a person with multimorbidity will be in separate stages concurrently, as they will be in a certain stage for each of their chronic condition. Moreover, given an ageing population and an increase in the numbers of PwCCs (and with multimorbidity) among them, the demand for chronic care will considerably increase worldwide [82]. Care models will no longer be able to rely solely on healthcare professionals because of lack of personnel and funding. Informal care will be essential for sustainability. These informal caregivers need to have the knowledge and be trained to provide care for the PwCC; at the same time, they also need to care for their own well-being. Supporting informal caregivers by equipping them with the knowledge to care for PwCC and for their own selves eventually will reflect on chronic care quality [30].

The PwCC journey relates well to the data collected from the scoping review, where aims and/or determinants and attributes of good quality chronic care have been applied and studied in the different stages of the journey (see supplementary file). This also strongly suggests that adopting what we propose to call as ‘journey consciousness’ should be expected from all actors (providers, users, regulators, financiers, policy-makers, etc.) who are in a position to shape the delivery of chronic care services.

### Aims

We identified seven aims for quality of chronic care in our scoping review: effectiveness; efficiency; safety; equity; accessibility, timeliness and affordability; person-centredness; and continuity. These are presented in our earlier paper [9].

### Determinants

We identified and classified ‘determinants’ of quality chronic care into our proposed four groups, which can be distributed across the different health system levels (national health system, local health system, health facility and healthcare team) akin to the macro-, meso- and micro-levels of the WHO ICCCF [68].

1. <u>Leadership & governance</u> would, at the national (systems) level, encompass the (overarching) policies and legislative frameworks for and in support of good quality care for chronic conditions. At the local and health facility levels, these would include different management and work systems, such as:

a. Local or health facility governance [19,32–33,38–41,43,48,50–51,54], local or health facility policies and local legislative frameworks as applicable (depending on the level of decentralization).
b. Quality management systems (QMS) [32–33,46,48–51,55].
c. Health information management systems [12–13,17–18,22–23,25,27,29,32–36,43,50,57–59].
d. Learning and knowledge management systems (including guidelines, standard operating procedures and their implementing rules and regulations) [14–17,22–23,25,28,32–33,43–45,47–49,51–52,54–55,57,59].
e. Resource governance / management of resources, including management of human resources for health [39–41,48,51–52,56].
f. Multisectoral and community engagement and collaborations to (help) address risks and determinants of chronic conditions, and to support the PwCC in their journey [20–21,25,26,33,36–39,50–52]. We note that stakeholders beyond the health sector are particularly relevant for chronic conditions, for co-designing and co-implementing interventions on risks and the social, structural and commercial determinants of the development and worsening of chronic conditions [33,38,39,43,49]. Such engagements can be initiated at both the national and local levels.
g. Involvement/engagement of PwCC in policy- and decision-making on issues concerning chronic care in general and for their own selves [20–21,25–26,33,36–39].
2. <u>Financing</u> at both national and local levels can consider healthcare financing itself, purchasing, healthcare spending (% of GDP, other sources, out-of-pocket expenditures), etc. Only five of the included scientific literature mention financing [25,32,43,46–47]. We note that three of our included literature suggest, but do not provide details, that rewards for effective clinical processes affecting management and prevention of chronic problems can be established[29,68,80]. This was one of the aspects explored in the second round of our Delphi survey and will be discussed in a separate paper.
3. <u>Resources</u> would depend on the level of (de)centralization of health care. For instance, in a (more) centralized system, most of these determinants would originate from the national level. Depending on the degree of decentralization, these would then shift towards the local health systems, and involve the health facilities, the communities, and the PwCC and their family. We identified determinants classified under resources from our scientific and grey literature, as follows:

a. Infrastructure – physical environment, healthcare facility buildings and lay-out of the buildings [60,65,81].
b. Health care staff – with skills, knowledge and expertise in caring for chronic conditions [52,56].
c. Health information – the information/data collected from people consulting at the health facility; what these are and how these are documented and kept, accessed, used, and shared [13–14,17–18,22–23,25,27,32–36,43,50,57–59].
d. Equipment, pharmaceuticals, diagnostics and consumables, based on local patterns of diseases and health problems, to support chronic care delivery from health promotion to tertiary prevention and to promote and support self-management and informal caregiver management of the PwCC [23,41,81].
e. Sectors and stakeholders other than health who may positively – directly or indirectly – contribute to prevention and control of risks and chronic conditions [33,38–40,43].
f. PwCC, their families and their social networks, and other resources in the community [15,17,22,32,43,50] noting that informal caregiver is the subject of one of the scientific literature we reviewed [30].
4. <u>Service delivery</u> This determinant group is well-developed in the included scientific literature, for instance:

a. chronic care services [5,12–81] including services for self-management education and support, and
b. delivery systems [27–29,32,34–40,42–43,46–52,54,56–60].

### Actions on the determinants

Corollary to the above, although the presence of some determinants *per se* can already be construed to (partly) achieve specific chronic care quality aims (e.g. having chronic care structures in place, i.e. availability of chronic care services), other determinants need particular actions.

As noted in the literature we reviewed, actions would include:

1. Organization of health services [12–13,15,17–20,22–25,30,32–34,36,39,43,46–47] to have a (comprehensive) offer of care for chronic conditions (i.e. the chronic care services). This includes sound determinations of what services should be offered and in what levels, which of the human resources for health are tasked to do which activity, and how these are organized, specifically (e.g. as a separate “vertical service” or integrated with other services). Organization should also include making the services available on a regular basis and considering financial, cultural and temporal accessibility. While certain services may not be available every day, the time schedule should be fixed (e.g. fixed hours and days) to facilitate access and maximally accommodate PwCCs.
2. Designing a system of delivery of chronic care that is conscious of a person’s journey through the natural history of chronic conditions, so that the offer is well-coordinated, continuous and seamless. A comprehensive health systems response would encompass health services from a person’s risk exposure to their end-of life. This way, PwCC do not fall from care as they traverse their journey and move from one level of care to another, and from the community to the health facilities and vice versa. We noted this journey-conscious action, describing specific steps in the journey (e.g., screening to diagnosis and follow-up; addressing complications and comorbidities and reintegration) in 15 of the scientific literature we reviewed [18,23–24,26–27,34,36–37,41,44,46,51,54,56,58].
3. Ensuring availability of appropriate equipment, laboratory tests and medicine responsive to the needs of the population and having proper inventory and stocking mechanisms to avoid stock-outs, mechanisms to avoid equipment breakdown [23,41,61,81].
4. Ensuring proper skill mix, training, professional education (i.e. updating knowledge with scientific evidence), and supportive supervision of human resources for health to deliver coordinated, collaborative, biopsychosocial, and person-centred, culture- and gender-sensitive chronic care including self-management education and support, to PwCC, as well as ensuring health care staff motivation [13,20,22,38–39,44,46–47,51,56–58].
5. Actions on health information systems so that:

a. Data are analysed in a timely manner to assess population needs [13,17,22,32,43,50]
b. Individual health records are available and up to date for individual case management [80]; and that said PwCC health record follow them from one service to another, from one level to another, and are also accessible to the PwCC (and their informal caregivers, as warranted) etc. [13–14,17–18,22–23,25,27,32–36,43,50,57–59].
6. Actions towards promoting a culture of quality such as continuous quality improvement, with a cycle of monitoring and evaluation to check for opportunities for improvement and to act on these constructively [33–34,44,47,49–52,56].
7. Actions on financing which would include, among others, proper allocation of finances; generation, mobilization, pooling and coordination of resources; incentives [25,32,38,67,75]; considerations for alternative financing mechanisms, etc. [24,26,33,35,39,47,49], considering that PwCCs are expected to utilize health services regularly throughout the duration of their condition and more often throughout their lifetime and, thus, would need and use resources more often.
8. Provision of PwCC self-management education and support (also for informal caregivers)[12–13,15–18,21–23,25,27,29–32,35,37,42–43,47,49,59].

### Integration as an exemplary principle to organize actions

Integration has received considerable attention and various interpretations, including in WHO [7,61–63,66]. Care and/or service integration is explicitly indicated as an action in 14 of the scientific literature we reviewed [18,22,33–35,37–38,40,43,45,55–57,59]. In 13 other reviewed scientific literature, the actions on care collaboration and/or coordination and/or partnerships were used as a means to integrate care or services or healthcare providers (both formal and informal) [16–17,22–24,36,39,42,44,46–47,52,54]. Our analysis places integration as an exemplary organising principle that orients actions on determinants to improve quality of chronic care. Organising the coherence of various interventions to improve quality of care matters, as acting on determinants presented above will also compete for resources.

Our literature review suggests that applying integration as an organizing principle to act on various chronic care quality determinants would have at least two implications.

First, it would encourage integration of different chronic care services, encompassing health promotion, and primary, secondary and tertiary prevention including curative care and involving different sectors and disciplines, and also taking into account social services [7,38–39,53,61,62,75,79,81]. This way, the different chronic care services at different levels and carried-out by various healthcare personnel/disciplines are *integrated* to: include population actions [25,33,38,56,59]; incorporate mechanisms to connect the PwCC with community resources [15,27,31,43,49,69,74]; and assure seamless access to all levels of care and/or other healthcare services/specialties (i.e. there is management continuity) and with their health information made available (i.e. informational continuity is ensured) covering all steps of the PwCC journey.

Second, it would encourage integration of ‘integrated chronic care services’ with other ongoing healthcare activities. Integration of diabetes care with other ongoing primary health care activities in LMICs has been demonstrated to improve quality of diabetes care in the past [83]. Community-based healthcare workers can perform health promotion, risk prevention and control, and screening for chronic conditions at the community level together with their other ongoing community-based activities (e.g. under-5 growth monitoring, as TB-DOTS treatment partner)[14,39]. Facility-based healthcare professionals can also take advantage and screen people identified at risk, if and when they consult (even for a different health problem), and confirm the diagnosis and initiate prompt treatment and self-management education of PwCC, together with their other healthcare activities[13].

### Attributes

We identified attributes relative to specific quality aims, per determinant group, from the included scientific and grey literature (Table 2). We note that these may be specific to the context of the setting/focus of the paper cited. While a considerable number of the scientific literature presented attributes of good clinical outcomes, we noted that attributes related to structure and/or process were also used.

**Table 1.**
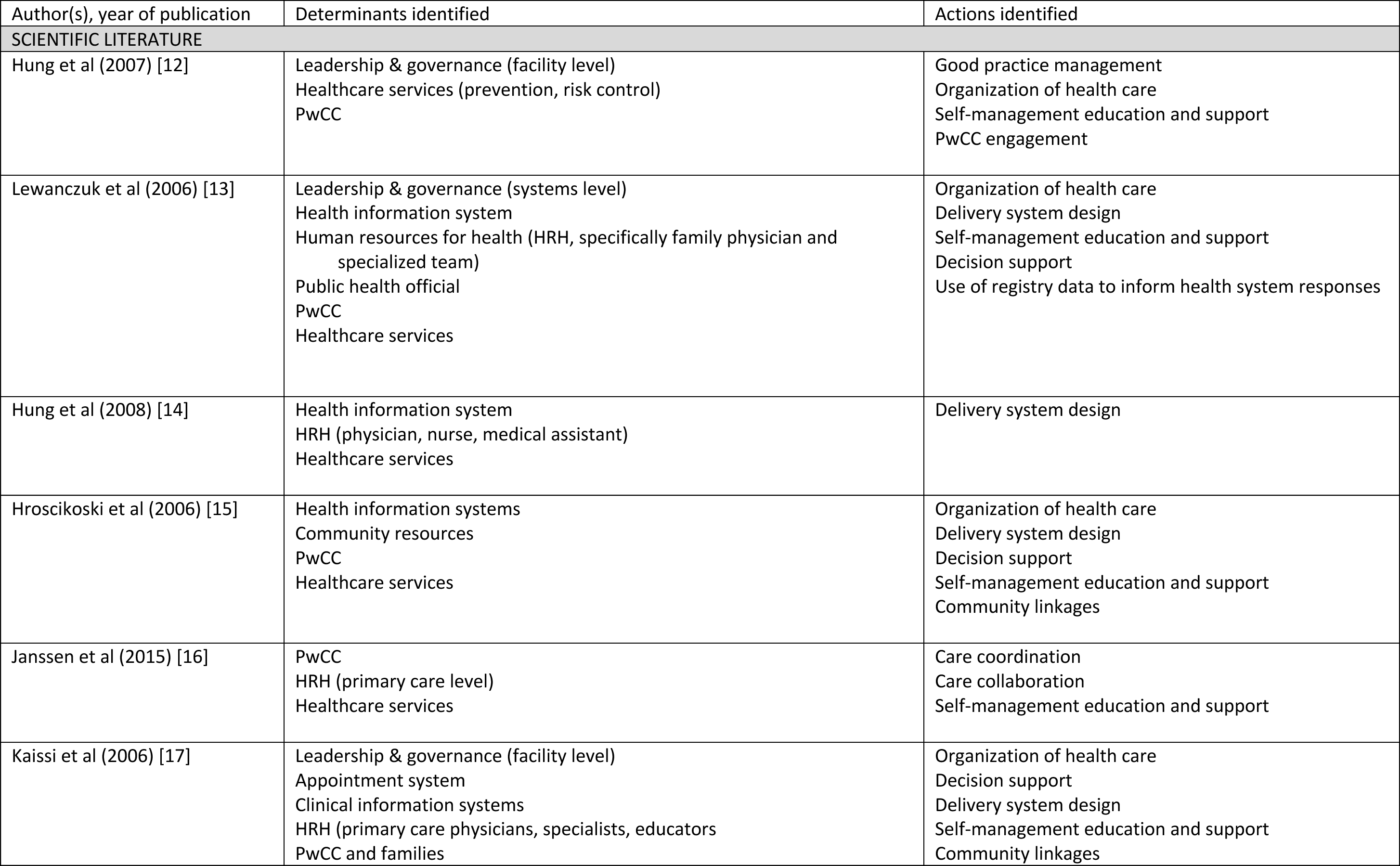
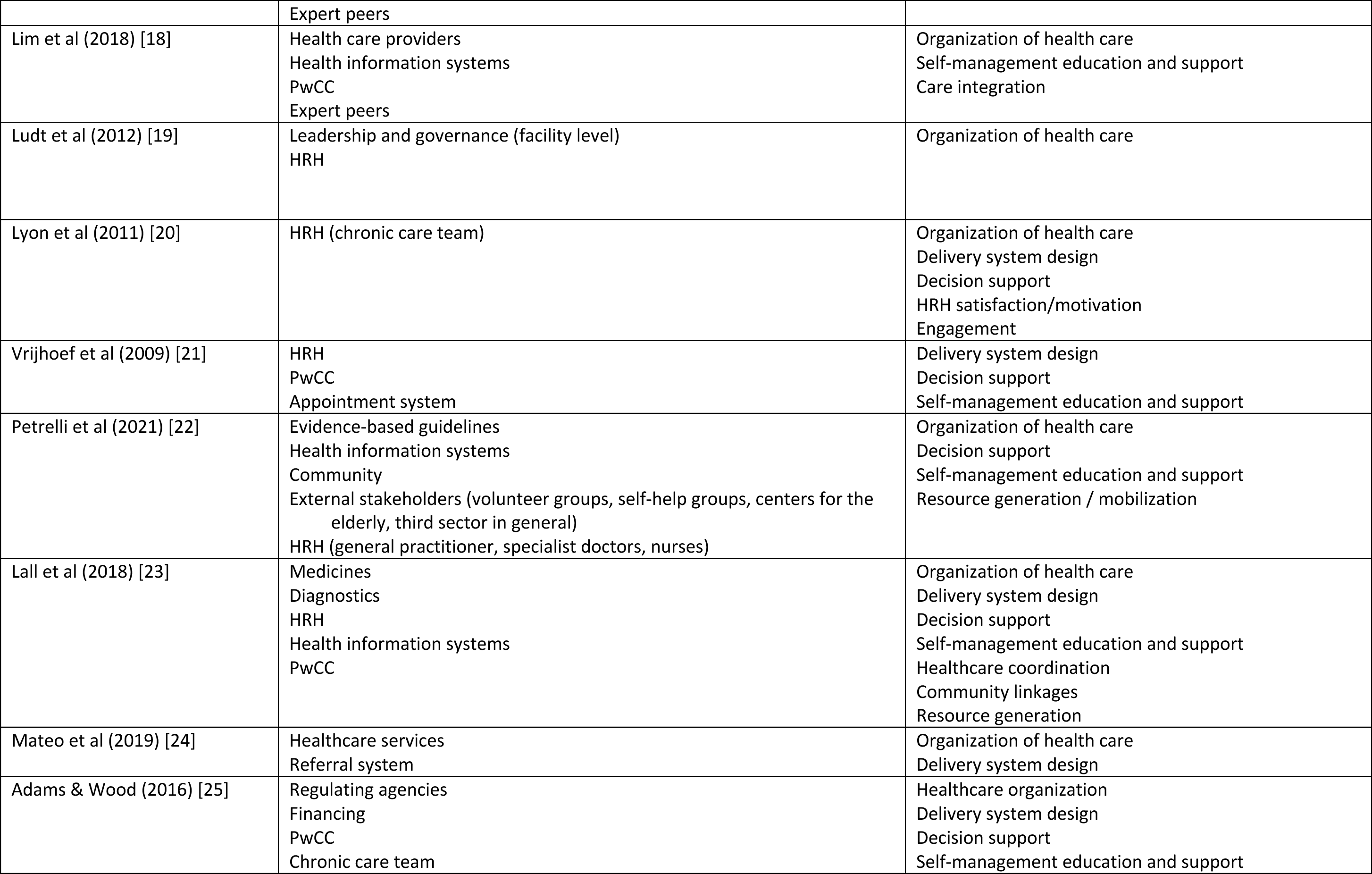
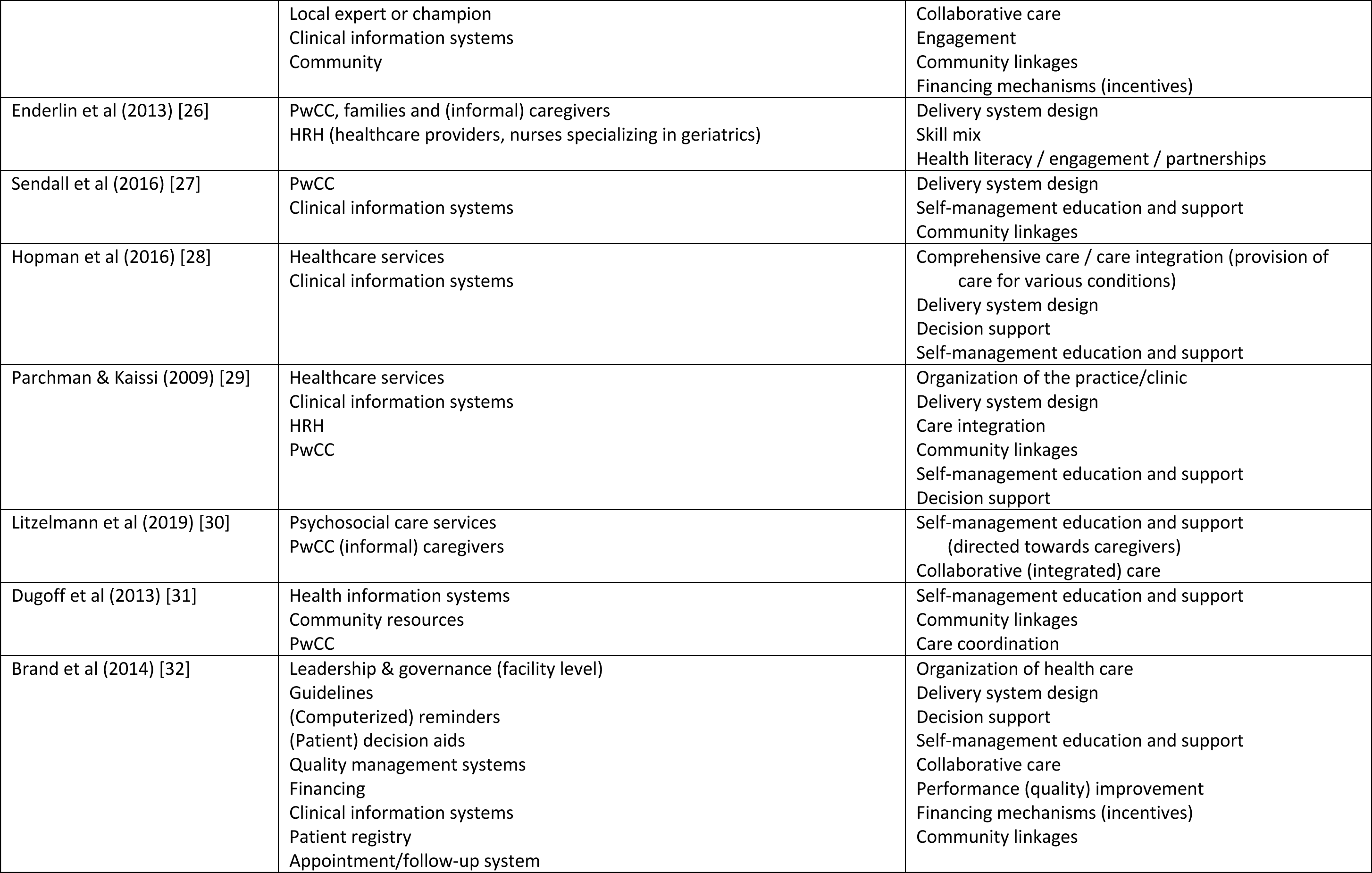
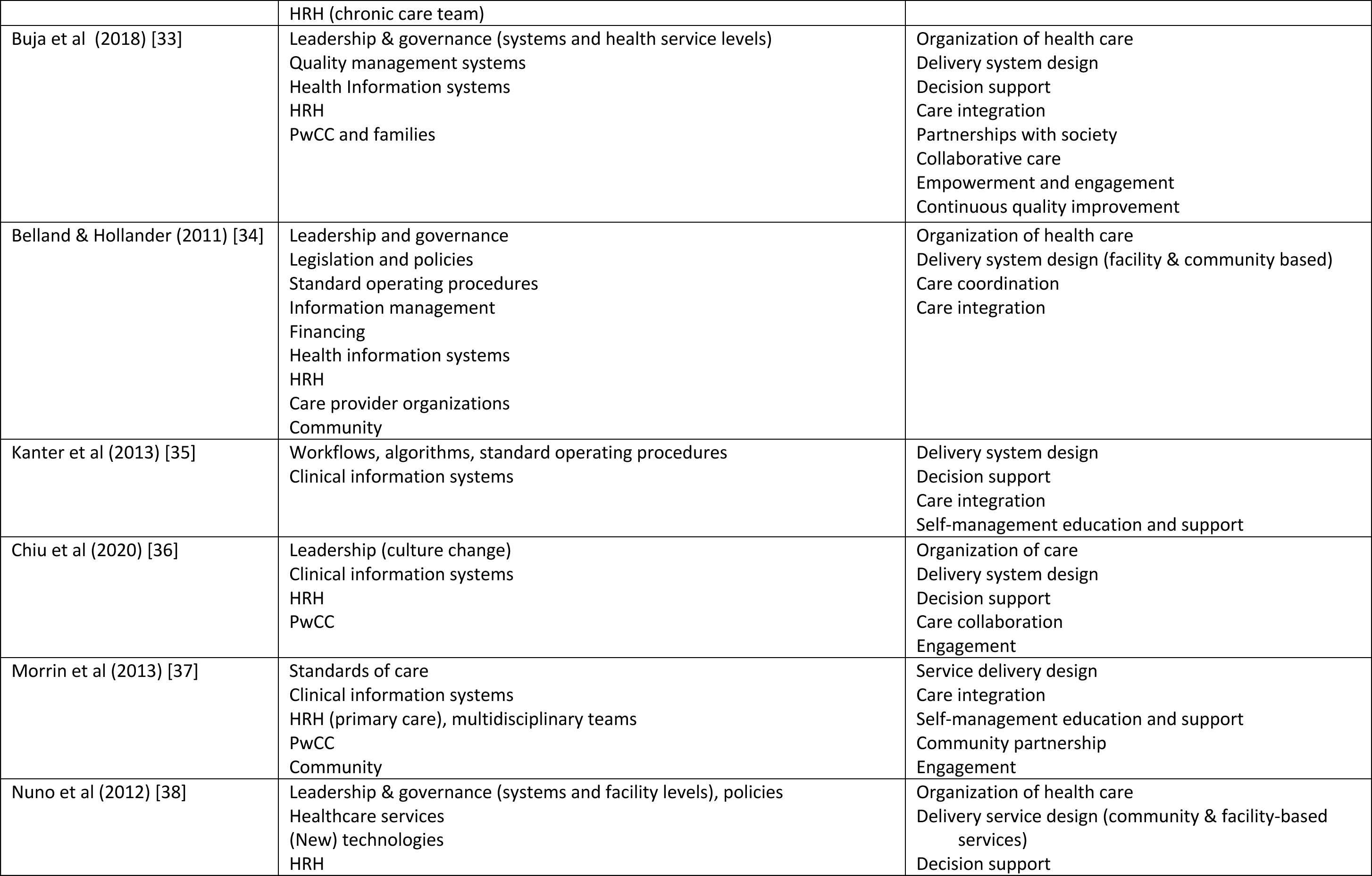
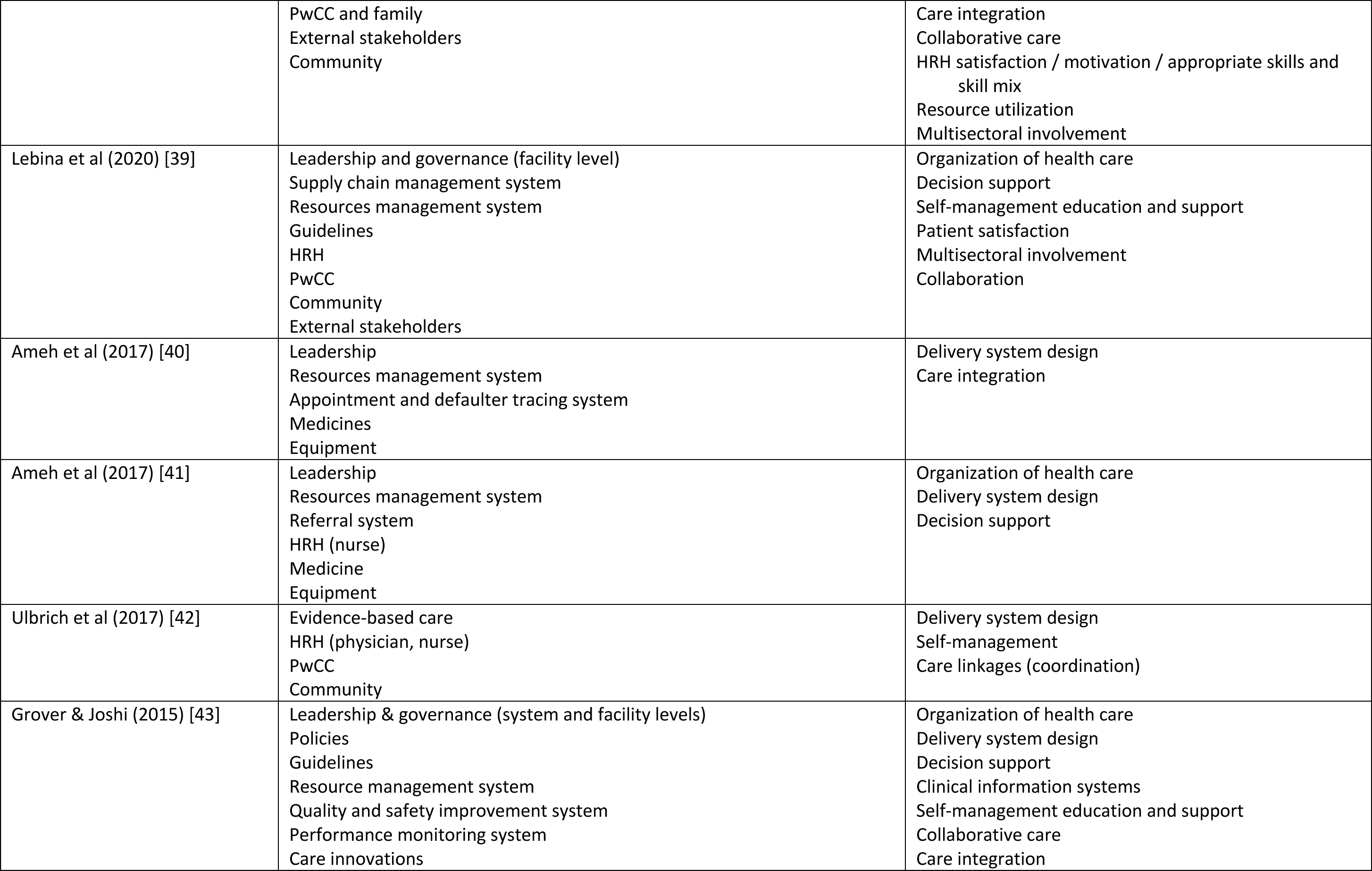
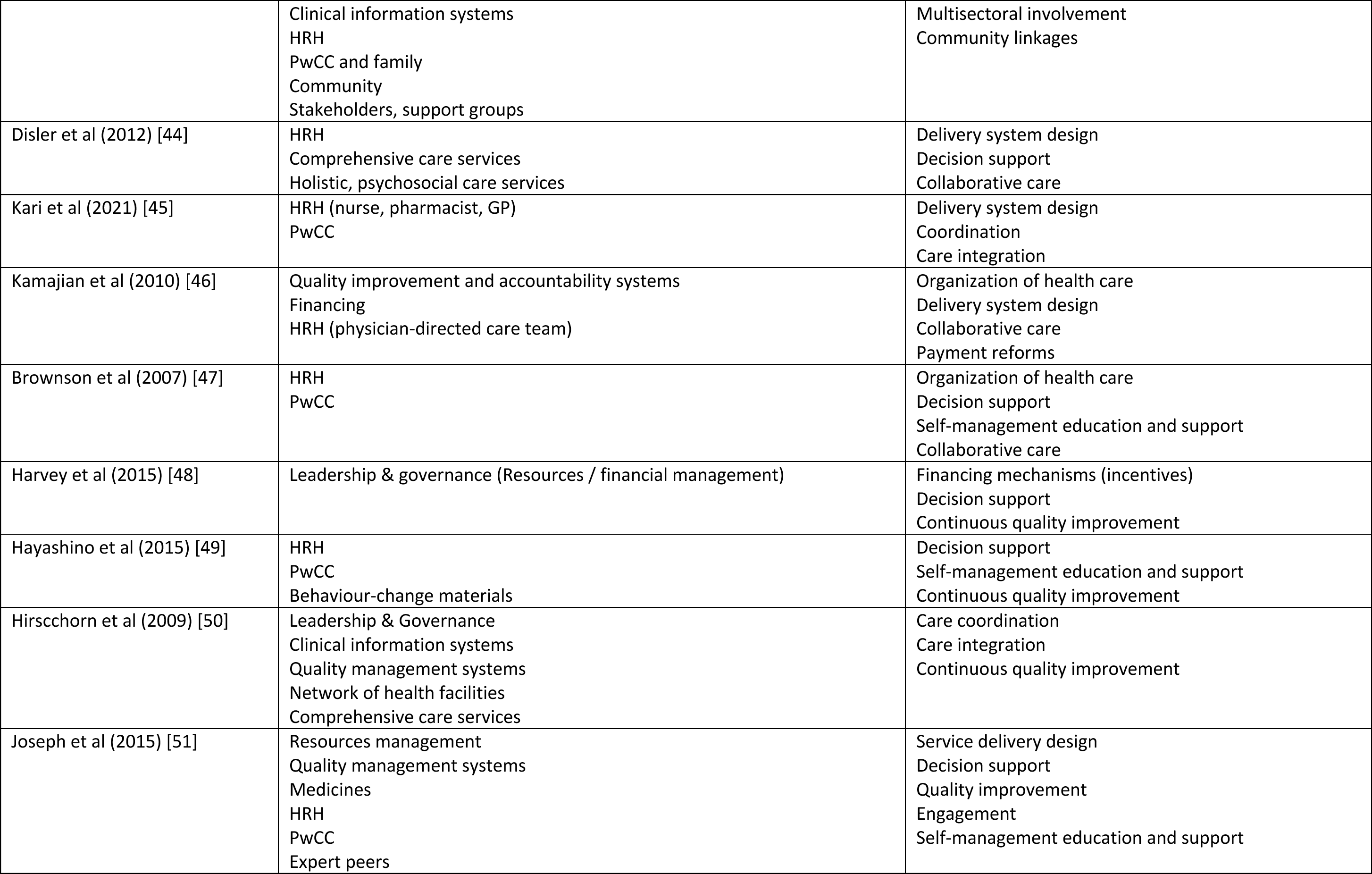
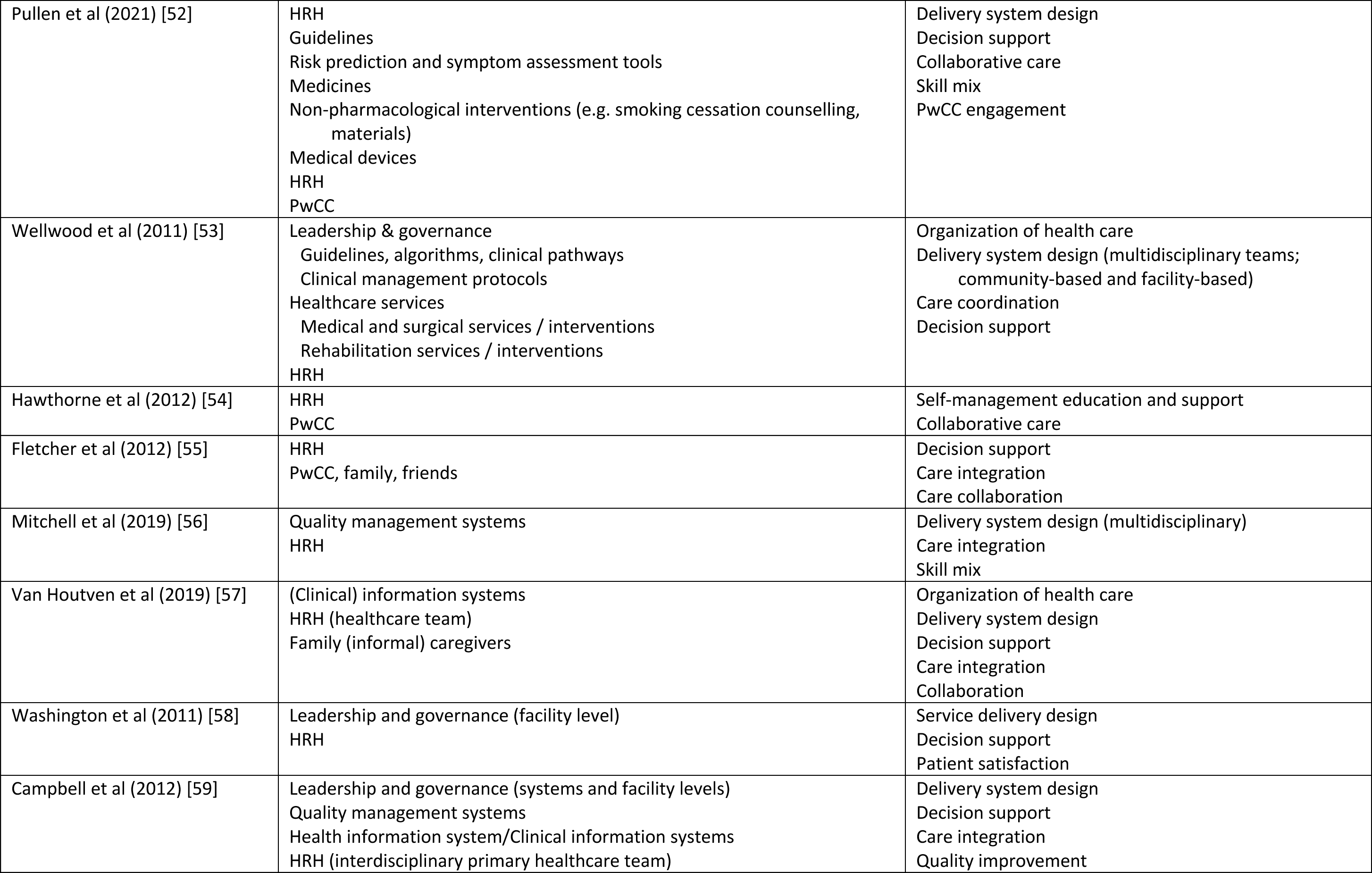
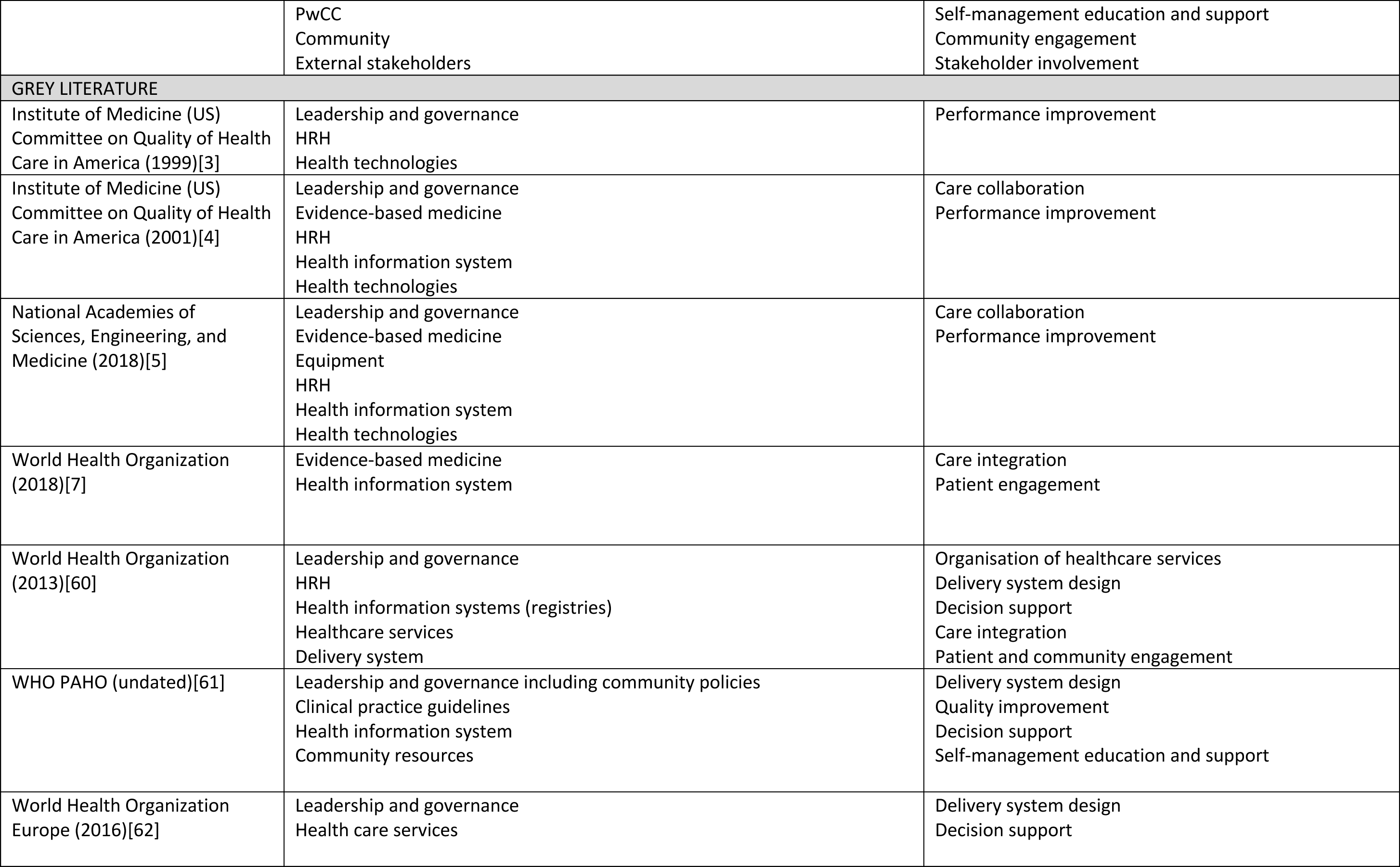
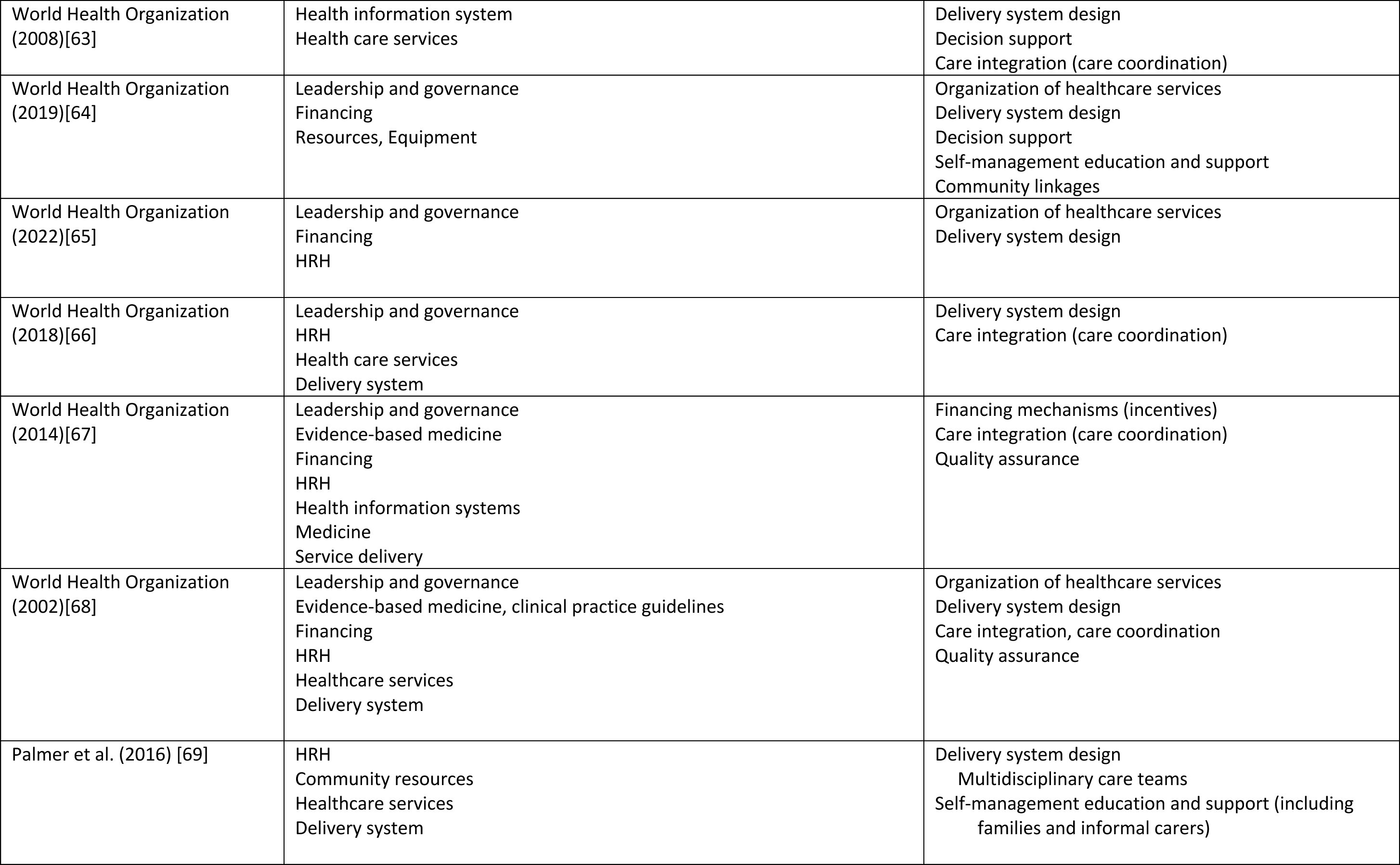
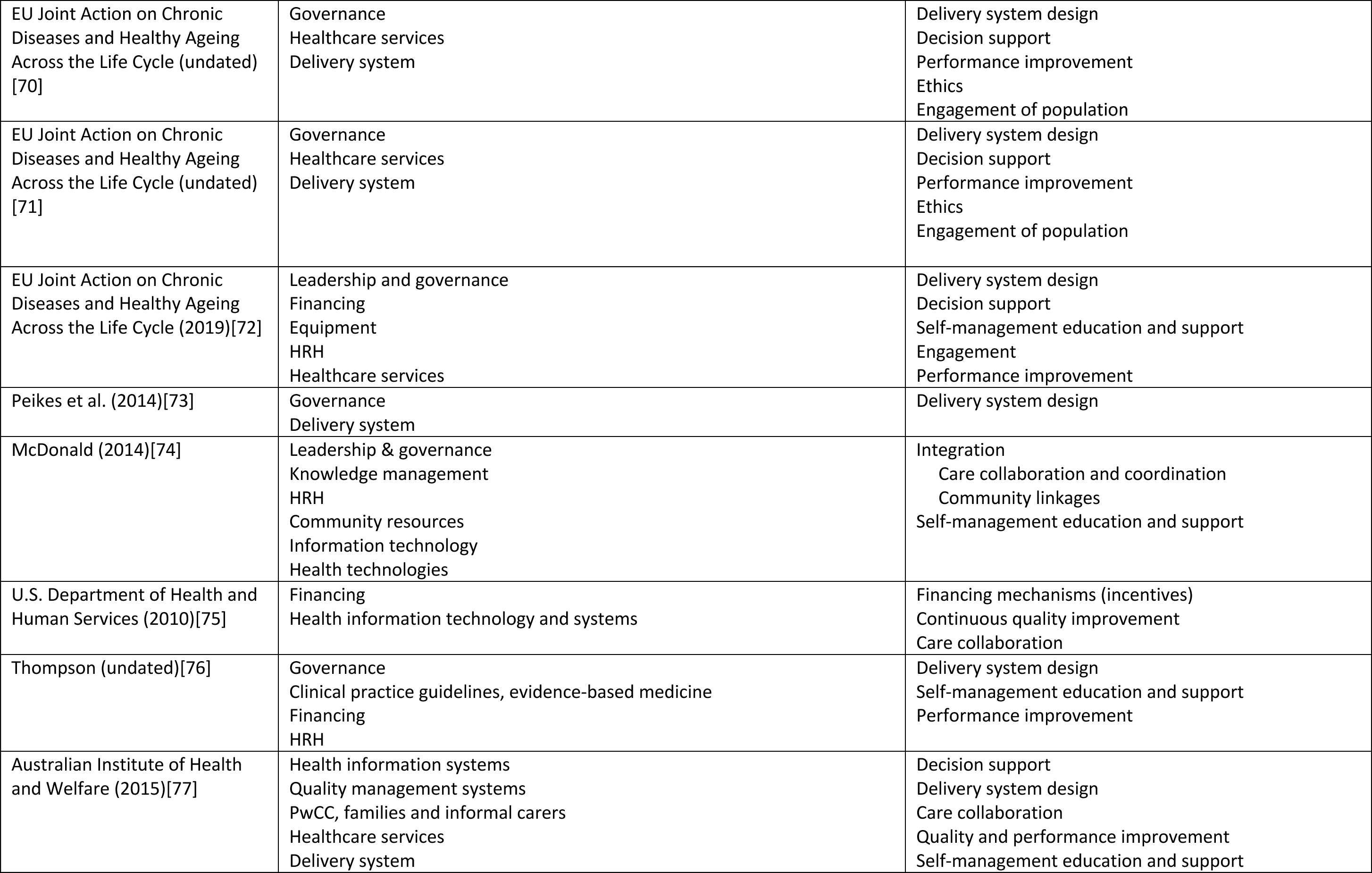
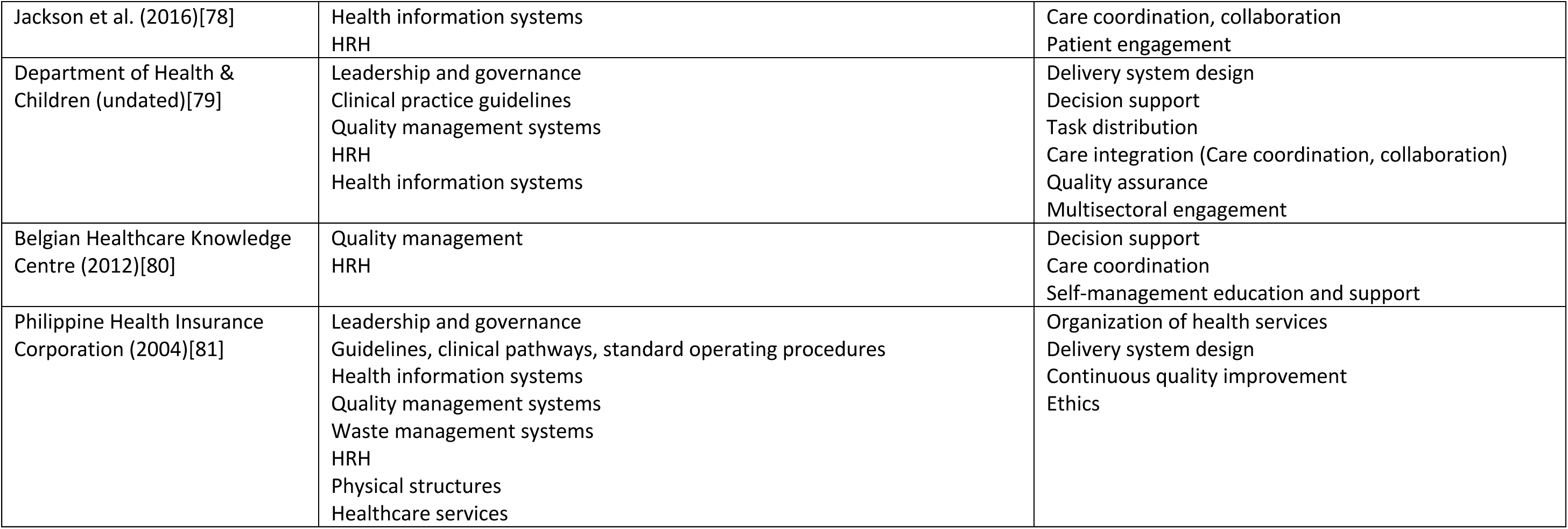
Chronic care quality determinants and actions identified from the literature reviewed.

| Author(s), year of publication | Determinants identified | Actions identified |
| --- | --- | --- |
| SCIENTIFIC LITERATURE |  |  |
| Hung et al (2007) [12] | Leadership & governance (facility level)<br>Healthcare services (prevention, risk control)<br>PwCC | Good practice management<br>Organization of health care<br>Self-management education and support<br>PwCC engagement |
| Lewanczuk et al (2006) [13] | Leadership & governance (systems level)<br>Health information system<br>Human resources for health (HRH, specifically family physician and specialized team)<br>Public health official<br>PwCC<br>Healthcare services | Organization of health care<br>Delivery system design<br>Self-management education and support<br>Decision support<br>Use of registry data to inform health system responses |
| Hung et al (2008) [14] | Health information system<br>HRH (physician, nurse, medical assistant)<br>Healthcare services | Delivery system design |
| Hroschikoski et al (2006) [15] | Health information systems<br>Community resources<br>PwCC<br>Healthcare services | Organization of health care<br>Delivery system design<br>Decision support<br>Self-management education and support<br>Community linkages |
| Janssen et al (2015) [16] | PwCC<br>HRH (primary care level)<br>Healthcare services | Care coordination<br>Care collaboration<br>Self-management education and support |
| Kaissi et al (2006) [17] | Leadership & governance (facility level)<br>Appointment system<br>Clinical information systems<br>HRH (primary care physicians, specialists, educators)<br>PwCC and families | Organization of health care<br>Decision support<br>Delivery system design<br>Self-management education and support<br>Community linkages |
|  | Expert peers |  |
| Lim et al (2018) [18] | Health care providers<br>Health information systems<br>PwCC<br>Expert peers | Organization of health care<br>Self-management education and support<br>Care integration |
| Ludt et al (2012) [19] | Leadership and governance (facility level)<br>HRH | Organization of health care |
| Lyon et al (2011) [20] | HRH (chronic care team) | Organization of health care<br>Delivery system design<br>Decision support<br>HRH satisfaction/motivation<br>Engagement |
| Vrijhoef et al (2009) [21] | HRH<br>PwCC<br>Appointment system | Delivery system design<br>Decision support<br>Self-management education and support |
| Petrelli et al (2021) [22] | Evidence-based guidelines<br>Health information systems<br>Community<br>External stakeholders (volunteer groups, self-help groups, centers for the elderly, third sector in general)<br>HRH (general practitioner, specialist doctors, nurses) | Organization of health care<br>Decision support<br>Self-management education and support<br>Resource generation / mobilization |
| Lall et al (2018) [23] | Medicines<br>Diagnostics<br>HRH<br>Health information systems<br>PwCC | Organization of health care<br>Delivery system design<br>Decision support<br>Self-management education and support<br>Healthcare coordination<br>Community linkages<br>Resource generation |
| Mateo et al (2019) [24] | Healthcare services<br>Referral system | Organization of health care<br>Delivery system design |
| Adams & Wood (2016) [25] | Regulating agencies<br>Financing<br>PwCC<br>Chronic care team | Healthcare organization<br>Delivery system design<br>Decision support<br>Self-management education and support |
|  | Local expert or champion<br>Clinical information systems<br>Community | Collaborative care<br>Engagement<br>Community linkages<br>Financing mechanisms (incentives) |
| Enderlin et al (2013) [26] | PwCC, families and (informal) caregivers<br>HRH (healthcare providers, nurses specializing in geriatrics) | Delivery system design<br>Skill mix<br>Health literacy / engagement / partnerships |
| Sendall et al (2016) [27] | PwCC<br>Clinical information systems | Delivery system design<br>Self-management education and support<br>Community linkages |
| Hopman et al (2016) [28] | Healthcare services<br>Clinical information systems | Comprehensive care / care integration (provision of care for various conditions)<br>Delivery system design<br>Decision support<br>Self-management education and support |
| Parchman & Kaissi (2009) [29] | Healthcare services<br>Clinical information systems<br>HRH<br>PwCC | Organization of the practice/clinic<br>Delivery system design<br>Care integration<br>Community linkages<br>Self-management education and support<br>Decision support |
| Litzelmann et al (2019) [30] | Psychosocial care services<br>PwCC (informal) caregivers | Self-management education and support (directed towards caregivers)<br>Collaborative (integrated) care |
| Dugoff et al (2013) [31] | Health information systems<br>Community resources<br>PwCC | Self-management education and support<br>Community linkages<br>Care coordination |
| Brand et al (2014) [32] | Leadership & governance (facility level)<br>Guidelines<br>(Computerized) reminders<br>(Patient) decision aids<br>Quality management systems<br>Financing<br>Clinical information systems<br>Patient registry<br>Appointment/follow-up system | Organization of health care<br>Delivery system design<br>Decision support<br>Self-management education and support<br>Collaborative care<br>Performance (quality) improvement<br>Financing mechanisms (incentives)<br>Community linkages |
|  | HRH (chronic care team) |  |
| Buja et al (2018) [33] | Leadership & governance (systems and health service levels)<br>Quality management systems<br>Health Information systems<br>HRH<br>PwCC and families | Organization of health care<br>Delivery system design<br>Decision support<br>Care integration<br>Partnerships with society<br>Collaborative care<br>Empowerment and engagement<br>Continuous quality improvement |
| Belland & Hollander (2011) [34] | Leadership and governance<br>Legislation and policies<br>Standard operating procedures<br>Information management<br>Financing<br>Health information systems<br>HRH<br>Care provider organizations<br>Community | Organization of health care<br>Delivery system design (facility & community based)<br>Care coordination<br>Care integration |
| Kanter et al (2013) [35] | Workflows, algorithms, standard operating procedures<br>Clinical information systems | Delivery system design<br>Decision support<br>Care integration<br>Self-management education and support |
| Chiu et al (2020) [36] | Leadership (culture change)<br>Clinical information systems<br>HRH<br>PwCC | Organization of care<br>Delivery system design<br>Decision support<br>Care collaboration<br>Engagement |
| Morrin et al (2013) [37] | Standards of care<br>Clinical information systems<br>HRH (primary care), multidisciplinary teams<br>PwCC<br>Community | Service delivery design<br>Care integration<br>Self-management education and support<br>Community partnership<br>Engagement |
| Nuno et al (2012) [38] | Leadership & governance (systems and facility levels), policies<br>Healthcare services<br>(New) technologies<br>HRH | Organization of health care<br>Delivery service design (community & facility-based services)<br>Decision support |
|  | PwCC and family<br>External stakeholders<br>Community | Care integration<br>Collaborative care<br>HRH satisfaction / motivation / appropriate skills and skill mix<br>Resource utilization<br>Multisectoral involvement |
| Lebina et al (2020) [39] | Leadership and governance (facility level)<br>Supply chain management system<br>Resources management system<br>Guidelines<br>HRH<br>PwCC<br>Community<br>External stakeholders | Organization of health care<br>Decision support<br>Self-management education and support<br>Patient satisfaction<br>Multisectoral involvement<br>Collaboration |
| Ameh et al (2017) [40] | Leadership<br>Resources management system<br>Appointment and defaulter tracing system<br>Medicines<br>Equipment | Delivery system design<br>Care integration |
| Ameh et al (2017) [41] | Leadership<br>Resources management system<br>Referral system<br>HRH (nurse)<br>Medicine<br>Equipment | Organization of health care<br>Delivery system design<br>Decision support |
| Ulbrich et al (2017) [42] | Evidence-based care<br>HRH (physician, nurse)<br>PwCC<br>Community | Delivery system design<br>Self-management<br>Care linkages (coordination) |
| Grover & Joshi (2015) [43] | Leadership & governance (system and facility levels)<br>Policies<br>Guidelines<br>Resource management system<br>Quality and safety improvement system<br>Performance monitoring system<br>Care innovations | Organization of health care<br>Delivery system design<br>Decision support<br>Clinical information systems<br>Self-management education and support<br>Collaborative care<br>Care integration |
|  | Clinical information systems<br>HRH<br>PwCC and family<br>Community<br>Stakeholders, support groups | Multisectoral involvement<br>Community linkages |
| Disler et al (2012) [44] | HRH<br>Comprehensive care services<br>Holistic, psychosocial care services | Delivery system design<br>Decision support<br>Collaborative care |
| Kari et al (2021) [45] | HRH (nurse, pharmacist, GP)<br>PwCC | Delivery system design<br>Coordination<br>Care integration |
| Kamajian et al (2010) [46] | Quality improvement and accountability systems<br>Financing<br>HRH (physician-directed care team) | Organization of health care<br>Delivery system design<br>Collaborative care<br>Payment reforms |
| Brownson et al (2007) [47] | HRH<br>PwCC | Organization of health care<br>Decision support<br>Self-management education and support<br>Collaborative care |
| Harvey et al (2015) [48] | Leadership & governance (Resources / financial management) | Financing mechanisms (incentives)<br>Decision support<br>Continuous quality improvement |
| Hayashino et al (2015) [49] | HRH<br>PwCC<br>Behaviour-change materials | Decision support<br>Self-management education and support<br>Continuous quality improvement |
| Hirschhorn et al (2009) [50] | Leadership & Governance<br>Clinical information systems<br>Quality management systems<br>Network of health facilities<br>Comprehensive care services | Care coordination<br>Care integration<br>Continuous quality improvement |
| Joseph et al (2015) [51] | Resources management<br>Quality management systems<br>Medicines<br>HRH<br>PwCC<br>Expert peers | Service delivery design<br>Decision support<br>Quality improvement<br>Engagement<br>Self-management education and support |
| Pullen et al (2021) [52] | HRH<br>Guidelines<br>Risk prediction and symptom assessment tools<br>Medicines<br>Non-pharmacological interventions (e.g. smoking cessation counselling, materials)<br>Medical devices<br>HRH<br>PwCC | Delivery system design<br>Decision support<br>Collaborative care<br>Skill mix<br>PwCC engagement |
| Wellwood et al (2011) [53] | Leadership & governance<br>Guidelines, algorithms, clinical pathways<br>Clinical management protocols<br>Healthcare services<br>Medical and surgical services / interventions<br>Rehabilitation services / interventions<br>HRH | Organization of health care<br>Delivery system design (multidisciplinary teams; community-based and facility-based)<br>Care coordination<br>Decision support |
| Hawthorne et al (2012) [54] | HRH<br>PwCC | Self-management education and support<br>Collaborative care |
| Fletcher et al (2012) [55] | HRH<br>PwCC, family, friends | Decision support<br>Care integration<br>Care collaboration |
| Mitchell et al (2019) [56] | Quality management systems<br>HRH | Delivery system design (multidisciplinary)<br>Care integration<br>Skill mix |
| Van Houtven et al (2019) [57] | (Clinical) information systems<br>HRH (healthcare team)<br>Family (informal) caregivers | Organization of health care<br>Delivery system design<br>Decision support<br>Care integration<br>Collaboration |
| Washington et al (2011) [58] | Leadership and governance (facility level)<br>HRH | Service delivery design<br>Decision support<br>Patient satisfaction |
| Campbell et al (2012) [59] | Leadership and governance (systems and facility levels)<br>Quality management systems<br>Health information system/Clinical information systems<br>HRH (interdisciplinary primary healthcare team) | Delivery system design<br>Decision support<br>Care integration<br>Quality improvement |
|  | PwCC<br>Community<br>External stakeholders | Self-management education and support<br>Community engagement<br>Stakeholder involvement |
| GREY LITERATURE |  |  |
| Institute of Medicine (US)<br>Committee on Quality of Health<br>Care in America (1999)[3] | Leadership and governance<br>HRH<br>Health technologies | Performance improvement |
| Institute of Medicine (US)<br>Committee on Quality of Health<br>Care in America (2001)[4] | Leadership and governance<br>Evidence-based medicine<br>HRH<br>Health information system<br>Health technologies | Care collaboration<br>Performance improvement |
| National Academies of<br>Sciences, Engineering, and<br>Medicine (2018)[5] | Leadership and governance<br>Evidence-based medicine<br>Equipment<br>HRH<br>Health information system<br>Health technologies | Care collaboration<br>Performance improvement |
| World Health Organization<br>(2018)[7] | Evidence-based medicine<br>Health information system | Care integration<br>Patient engagement |
| World Health Organization<br>(2013)[60] | Leadership and governance<br>HRH<br>Health information systems (registries)<br>Healthcare services<br>Delivery system | Organisation of healthcare services<br>Delivery system design<br>Decision support<br>Care integration<br>Patient and community engagement |
| WHO PAHO (undated)[61] | Leadership and governance including community policies<br>Clinical practice guidelines<br>Health information system<br>Community resources | Delivery system design<br>Quality improvement<br>Decision support<br>Self-management education and support |
| World Health Organization<br>Europe (2016)[62] | Leadership and governance<br>Health care services | Delivery system design<br>Decision support |
| World Health Organization (2008)[63] | Health information system<br>Health care services | Delivery system design<br>Decision support<br>Care integration (care coordination) |
| World Health Organization (2019)[64] | Leadership and governance<br>Financing<br>Resources, Equipment | Organization of healthcare services<br>Delivery system design<br>Decision support<br>Self-management education and support<br>Community linkages |
| World Health Organization (2022)[65] | Leadership and governance<br>Financing<br>HRH | Organization of healthcare services<br>Delivery system design |
| World Health Organization (2018)[66] | Leadership and governance<br>HRH<br>Health care services<br>Delivery system | Delivery system design<br>Care integration (care coordination) |
| World Health Organization (2014)[67] | Leadership and governance<br>Evidence-based medicine<br>Financing<br>HRH<br>Health information systems<br>Medicine<br>Service delivery | Financing mechanisms (incentives)<br>Care integration (care coordination)<br>Quality assurance |
| World Health Organization (2002)[68] | Leadership and governance<br>Evidence-based medicine, clinical practice guidelines<br>Financing<br>HRH<br>Healthcare services<br>Delivery system | Organization of healthcare services<br>Delivery system design<br>Care integration, care coordination<br>Quality assurance |
| Palmer et al. (2016) [69] | HRH<br>Community resources<br>Healthcare services<br>Delivery system | Delivery system design<br>Multidisciplinary care teams<br>Self-management education and support (including families and informal carers) |
| EU Joint Action on Chronic Diseases and Healthy Ageing Across the Life Cycle (undated) [70] | Governance<br>Healthcare services<br>Delivery system | Delivery system design<br>Decision support<br>Performance improvement<br>Ethics<br>Engagement of population |
| EU Joint Action on Chronic Diseases and Healthy Ageing Across the Life Cycle (undated) [71] | Governance<br>Healthcare services<br>Delivery system | Delivery system design<br>Decision support<br>Performance improvement<br>Ethics<br>Engagement of population |
| EU Joint Action on Chronic Diseases and Healthy Ageing Across the Life Cycle (2019)[72] | Leadership and governance<br>Financing<br>Equipment<br>HRH<br>Healthcare services | Delivery system design<br>Decision support<br>Self-management education and support<br>Engagement<br>Performance improvement |
| Peikes et al. (2014)[73] | Governance<br>Delivery system | Delivery system design |
| McDonald (2014)[74] | Leadership & governance<br>Knowledge management<br>HRH<br>Community resources<br>Information technology<br>Health technologies | Integration<br>Care collaboration and coordination<br>Community linkages<br>Self-management education and support |
| U.S. Department of Health and Human Services (2010)[75] | Financing<br>Health information technology and systems | Financing mechanisms (incentives)<br>Continuous quality improvement<br>Care collaboration |
| Thompson (undated)[76] | Governance<br>Clinical practice guidelines, evidence-based medicine<br>Financing<br>HRH | Delivery system design<br>Self-management education and support<br>Performance improvement |
| Australian Institute of Health and Welfare (2015)[77] | Health information systems<br>Quality management systems<br>PwCC, families and informal carers<br>Healthcare services<br>Delivery system | Decision support<br>Delivery system design<br>Care collaboration<br>Quality and performance improvement<br>Self-management education and support |
| Jackson et al. (2016)[78] | Health information systems<br>HRH | Care coordination, collaboration<br>Patient engagement |
| Department of Health & Children (undated)[79] | Leadership and governance<br>Clinical practice guidelines<br>Quality management systems<br>HRH<br>Health information systems | Delivery system design<br>Decision support<br>Task distribution<br>Care integration (Care coordination, collaboration)<br>Quality assurance<br>Multisectoral engagement |
| Belgian Healthcare Knowledge Centre (2012)[80] | Quality management<br>HRH | Decision support<br>Care coordination<br>Self-management education and support |
| Philippine Health Insurance Corporation (2004)[81] | Leadership and governance<br>Guidelines, clinical pathways, standard operating procedures<br>Health information systems<br>Quality management systems<br>Waste management systems<br>HRH<br>Physical structures<br>Healthcare services | Organization of health services<br>Delivery system design<br>Continuous quality improvement<br>Ethics |

**Table 2.** (Some) attributes of the chronic care quality dimensions, per determinant group and considering the PwCC journey through healthcare, as determined from the scoping review and Delphi survey.

| Quality Aim | Leadership & Governance | Financing | Resources | Service Delivery |
| --- | --- | --- | --- | --- |
| **Effectiveness** | Relevant policies and monitoring and evaluation of implementation [38,43].<br>Availability and usage of clinical practice guidelines and standard operating procedures. | Proper resource allocation to prioritize certain health services, to ensure delivery of effective services and to ensure adequate population coverage [68]. | Temperature, air quality, lighting and noise conditions are within acceptable levels to maintain staff well-being and enhance effective work outcomes[5].<br>HCPs are trained regarding risks and determinants of chronic conditions | Services offered respond to the needs of the population and PwCCs.<br>Services offered follow guidelines and SoPs [17,29,32,39,52]. |
|  | Continuous quality improvement is in place. Supportive supervision is practiced.<br>Financial management is in place to ensure availability of sufficient resources to meet healthcare needs [65].<br>Health management information systems are in place and data analysed in a timely manner to determine the needs of the population and PwCCs [33,55] and used in the management planning of health services and resources[13].<br>Human resources management is in place to ensure availability of health care staff with skills, knowledge and expertise in caring for chronic conditions and the proper “skill mix”[56].<br>M&E of effectiveness of care delivered (e.g., clinical audits, mortality conferences)[43,71]. |  | (health promotion & prevention); to deliver chronic care (early diagnosis, follow-up, transition / reintegration); and to provide (self) management education and support to PwCC and informal caregiver[13,15,22].<br>Health information are recorded effectively[75]. | Care delivered is congruent to people’s (diverse) beliefs and values (cultural effectiveness)[71].<br>Care for chronic conditions effectively decreases the numbers of chronic conditions and complications and prevents and relieves suffering, through:<br>• Identification of those at risk and limiting said risks for developing chronic conditions[12,65].<br>• Early diagnosis and effective, prompt and regular treatment [52] of the chronic condition, both pharmacologically & non-pharmacologically (technical effectiveness [49].<br>• Counseling (psychological support, dietary, healthy lifestyle)[19].<br>• Co-management of comorbidities / multimorbidities [69] (both acute and chronic) and other health problems.<br>• Prevention of emergence of complications to the extent possible[18,25].<br>• Provision of appropriate palliative and end-of-life care[36,44]<br>• Proper care coordination and linking to care[50], including<br>self- and informal caregiver management[27]. |
| **Efficiency** | No evidences of poor management, fraud, corruption, and abusive practices[5]. | Cost-efficiency of health services[32]. | Members of the healthcare team are familiar with one another and work well together to provide the highest level of care in the most efficient manner[74]. Staff and PwCC have proper forum to express ideas on healthcare and its delivery[33]. | Acute and chronic care services are integrated to provide efficiency of healthcare service delivery[27]. |
| **Safety** | Building and physical environment management is in place[81]. Clinical risk management to prevent and control adverse care incidents [33] and effectively and efficiently address adverse effects [18] is in place. | Proper resource allocation to maintain building and physical environment management, clinical risk management, and care coordination and communications [65,68] | Air quality and noise conditions are within acceptable limits[33]. Building infrastructure is friendly to people with physical disabilities. Infrastructure is well-equipped to prevent falls. Proper sanitation, hygiene and (hazardous) waste management are maintained[81]. Essential equipment needed to provide safe essential health services are available and functioning [65]. Prevention of breakdowns in communication and care coordination across settings and healthcare providers[19]. | Care delivered in different levels of healthcare and related services including in the community is consistent[26]. |
| **Equity** | Resource pooling and purchasing functions of financial management are in place to assure equitable access and get resources to them who need it most[65]. Linked to access: improved, equitable access to care for those with chronic conditions is most likely to occur with multiple linked strategies that target different levels of the health system, and availability[53] and affordability of services[23,40]. | Out-of-pocket expenditures are reasonable and that no one is left behind / no one is pushed behind. Availability of alternative financing mechanisms in cases of limitations in regular financing[23]. | Healthcare staff have legitimacy [40] and are sensitive to beliefs, values, culture [43,74], ethnicity, disabilities, gender [58], etc of PwCC | Care delivered to all PwCCs is consistent[47]. |
| **Accessibility, timeliness and affordability** | Mechanisms are in place to ensure that resources are made available in a timely manner[65]. | Health financing strategies increase access to healthcare (make health services financially accessible / affordable to all)[40]. | Design of the physical structure ensures ease of access for all PwCCs.<br>Health care workers are accessible, e.g. culturally congruent, approachable[40]<br>Health information is recorded and shared in a timely manner to healthcare providers across disciplines and levels[42]. | Minimum basic services for chronic conditions providing a continuum of care [34] that include diagnosis, pharmacological and non-pharmacological treatment including counselling, self-management education and support, psychosocial support, targeted risk control and higher level of care (through an established referral system), etc. that are acceptable to the population served are available [43].<br>Services are temporally accessible (opening hours are accessible to people who utilise the services) and are available regularly with set schedules [23,41]. |
| **Person-centredness** | PwCCs work collaboratively with policy makers / health professionals / health managers in promoting and improving chronic care including palliative / end-of-life care[36,43]. | Finances are available to empower and engage PwCCs and to provide self-management education and support. | HCWs have skills and expertise on motivational interviewing, stages of change theory, goal setting, action plan development, patient self-management support [20] and in the delivery of holistic care taking into consideration not only the clinical aspects of the chronic condition but also the psychosocial aspects[7] of the PwCC. | The PwCC is involved in collaborative decision-making[25], and health empowerment / engagement is provided.<br>Care delivered also includes collaboration with informal caregivers[30].<br>Care delivered is congruent with the (diverse) beliefs, values and contexts of PwCC[32]. |
| **Continuity** | There is a systematic approach to planning and coordinating care | Funds are allocated for care planning, coordination, referral and | There is trust [41,45] and (good) rapport [19] between the PwCC (and/or the | The delivery of health promotion and all levels of prevention |
|  | [32] from controlling risks to end-of-life and including self-management (PwCC and informal caregiver). | counter-referral in all levels (community and facilities) and disciplines and including support to PwCC and informal caregivers for care continuum at home. | informal caregiver) and their healthcare providers (relational continuity). Communication and care coordination across settings and healthcare providers is established.[19]<br>Community resources are identified / mapped-out. | (primary, secondary, tertiary; community-based, primary healthcare facility-based, hospital-based) is well-coordinated and as seamless as possible, from the home, to the community to healthcare facilities, and transitioning from healthcare facilities to the community and the home[26,27], with special attention to comorbidities / multimorbidities and the elderly [26,41] with chronic conditions. For instance, clinical care pathways / case management plans[31], referral and counter-referral mechanisms across care disciplines and levels [53] (community, primary, secondary, tertiary levels) are in place and are implemented. |

### Delphi survey results

Forty-nine of the 52 invited stakeholders (94%) consented and participated in the Delphi survey. Table 3 provides demographic and pertinent characteristics.

**Table 3.**
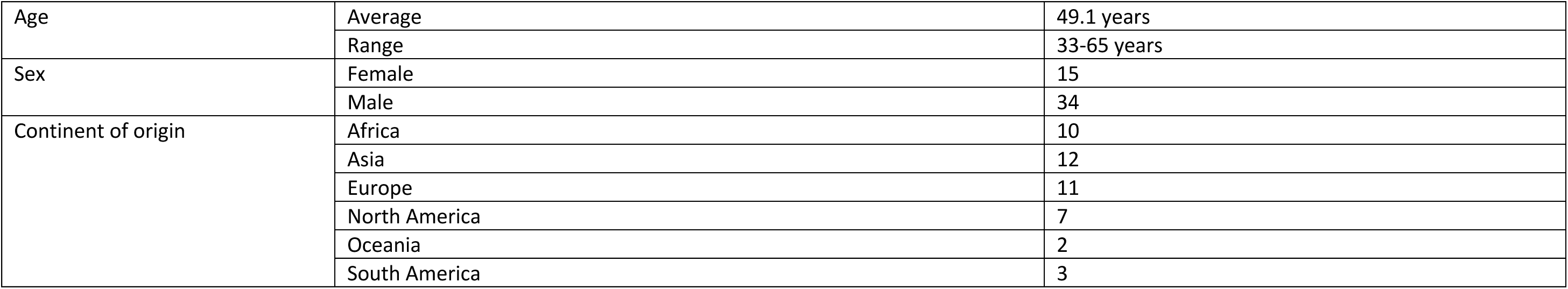
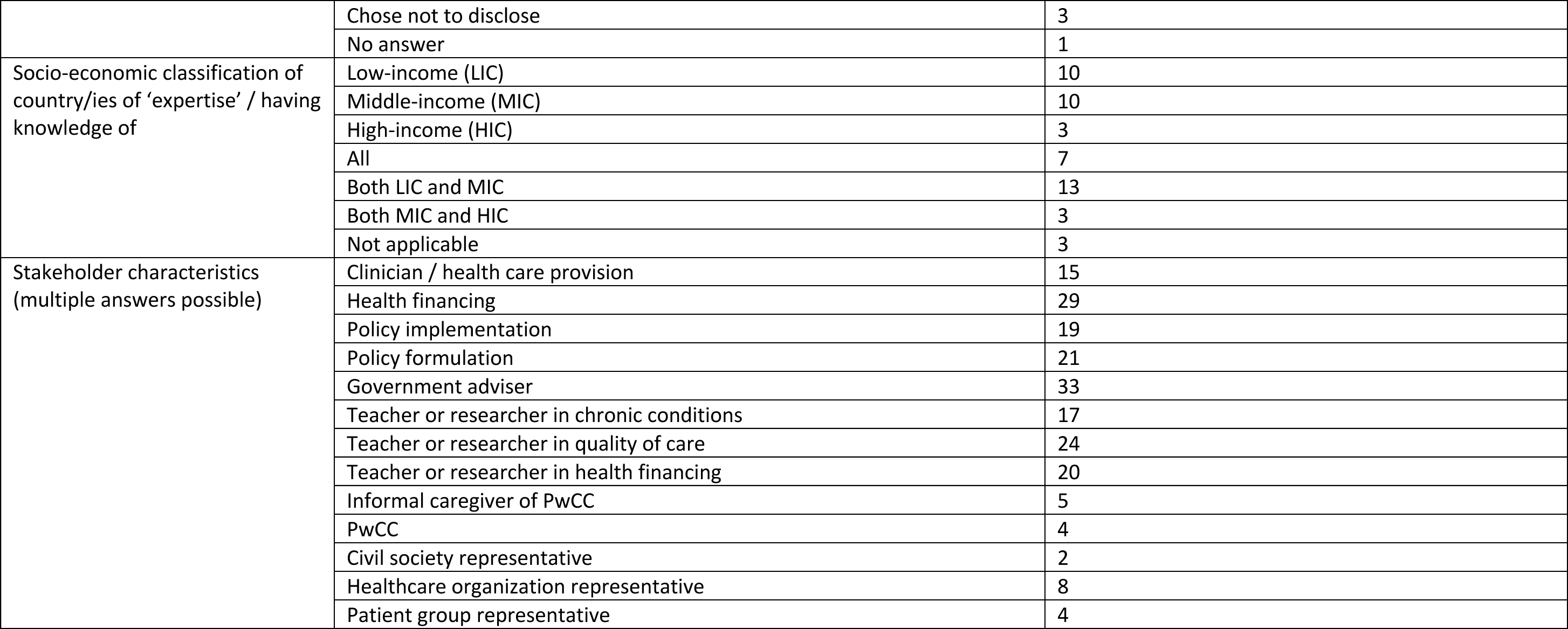
Demographic characteristics of Delphi respondents (n=49)

|  |  |  |
| --- | --- | --- |
| Age | Average | 49.1 years |
|  | Range | 33-65 years |
| Sex | Female | 15 |
|  | Male | 34 |
| Continent of origin | Africa | 10 |
|  | Asia | 12 |
|  | Europe | 11 |
|  | North America | 7 |
|  | Oceania | 2 |
|  | South America | 3 |
|  | Chose not to disclose | 3 |
|  | No answer | 1 |
| Socio-economic classification of country/ies of 'expertise' / having knowledge of | Low-income (LIC) | 10 |
|  | Middle-income (MIC) | 10 |
|  | High-income (HIC) | 3 |
|  | All | 7 |
|  | Both LIC and MIC | 13 |
|  | Both MIC and HIC | 3 |
|  | Not applicable | 3 |
| Stakeholder characteristics (multiple answers possible) | Clinician / health care provision | 15 |
|  | Health financing | 29 |
|  | Policy implementation | 19 |
|  | Policy formulation | 21 |
|  | Government adviser | 33 |
|  | Teacher or researcher in chronic conditions | 17 |
|  | Teacher or researcher in quality of care | 24 |
|  | Teacher or researcher in health financing | 20 |
|  | Informal caregiver of PwCC | 5 |
|  | PwCC | 4 |
|  | Civil society representative | 2 |
|  | Healthcare organization representative | 8 |
|  | Patient group representative | 4 |

There was consensus on the proposed chronic care quality aims, as we described previously [9]. In the open-ended component of the survey, the respondents described determinants and actions, and related these to achievement of the aims based on their experiences, more particularly in low resource settings. The panellists agree that attributes need to be contextual, and that the structure-process-outcome framework can be used to identify specific indicators.

They indicated the need to organize chronic care and service delivery that encompasses the whole gamut from screening to clinical management (or a ‘journey-conscious’ approach), which could redound on efficiency, accessibility, equity, effectiveness and continuity.

*“prevention must be considered”*
*“Different segments of the health system could provide better continuity of care, from screening through management through treatment - which would make a big difference to patients.”*
*“the passive and disease-focused approach of the healthcare system affects the quality of chronic care, which is often reduced to the belated attention of clinical complications in referral settings and at a high cost”*
*“Service delivery for chronic conditions is absent in primary care. Medicines are not continuously available or not affordable or it is not well explained how they must be taken or adhered to or people receive the inappropriate medicines for their condition.”*
*“Right now, we have reactive, short-term care and changing that mindset is priority. (Currently) chronic care is nobody’s business which, unfortunately, means that patients have to do it (by) themselves.”*

The need for resources was highlighted; specifically identifying human resources to improve accessibility and equity, having health information systems in place for care continuity, and engagement of PwCC for self-management.

*“provision of health care needs resources”*
*“Unequal distribution of doctors with concentrations in urban areas aggravates access problems.”*
*“I would ensure that services of health providers were generally accessible to all and equitably distributed.”*
*“…the absence of good electronic MIS (health information systems) that track the patient contributes to lack of continuity of care.”*
*“I would put a premium on empowering patients to be able to do self-care, particularly for interventions that have been proven to be effective. This means increasing health literacy and emphasis on primary prevention.”*

Regarding financing, the respondents indicated effects on accessibility, equity, continuity and person-centredness.

*“The greatest current challenge is the lack of financing… the health system was under-funded…accessibility worsened following the political-economic crisis. Inequity has always been a major challenge”*
*“Without a single financial architecture, the data systems, continuity of care, patient-centeredness, etc. are all extremely difficult to achieve.”*

The responses were varied for the determinant group leadership and governance, calling for strategic planning and priority setting, and connecting these determinants with integration, collaborative care and wider determinants of health, which would redound on chronic care quality:

*“Work more on strategic planning and setting priorities, as well as improving health care in general.”*
*“For improved care on chronic conditions we need to achieve the following:*

*a. Delivery within integrated care pathways spanning across care sectors and being organised around the patient;*
b. *Co-development of interventions, Care pathways and treatment plans with patients;*
c. *Provision of the data, electronic, administrative, and governance infrastructure to enable integrated care for chronic condition;*
d. *Focus on improving wider determinants of health (housing, social support network, community facilities, education etc) and primary prevention in the patient’s community setting.”*

They also mentioned strengthening the health information and referral and counter-referral systems to help ensure continuity:

*“(Continuity) can be enabled by strengthening the information system and introducing digitization, where possible.”*
*“we need to set up a good health information system to monitor patients within the district. Depending on the technical facilities available, a referral and counter-referral system needs to be set up …between the 2 levels … to deal with any complications.”*

and working on human resources for health to improve retention.

*“improve the plans for health staff and develop motivational mechanisms for keeping health staff in a country.”*

For our proposed framework, the panel recommended viewing equity as a cross-cutting issue, which calls for broad societal transformations. Inequitable access to care for chronic conditions is evident: while the better-off and urban population can avail private facilities or outpatient care at hospital level, the poorest households will often not find accessible or affordable treatments in their surroundings. This calls for attention to vulnerable groups (e.g. migrants, ethnic minorities, marginalized population, people living with disability), the extra barriers they face (e.g. cultural barriers, stigma, etc.) and the lower quality of care they often get.

Considering the serious shortcomings in quality of care in many facilities and health systems, the panel recommended prioritization to ensure that core determinants of quality of care are fulfilled (i.e. secure the availability of medicines, diagnostics, qualified staff, appropriate digital data system, quality assurance and clinical governance, etc.).

### Illustrating the chronic care quality framework

We finalised the chronic care quality framework considering the the above results.

Figure 4 illustrates that actions on determinants can fulfil quality aims and any of these can be measured through attributes, making use of structure, process and outcomes criteria and indicators. We also emphasized the view that equity should be a cross-cutting aim, as recommended by the Delphi panel. We remind that the determinants and actions should be situated in each step of the healthcare journey of PwCC, as illustrated in Figure 3.

**Figure 4.**
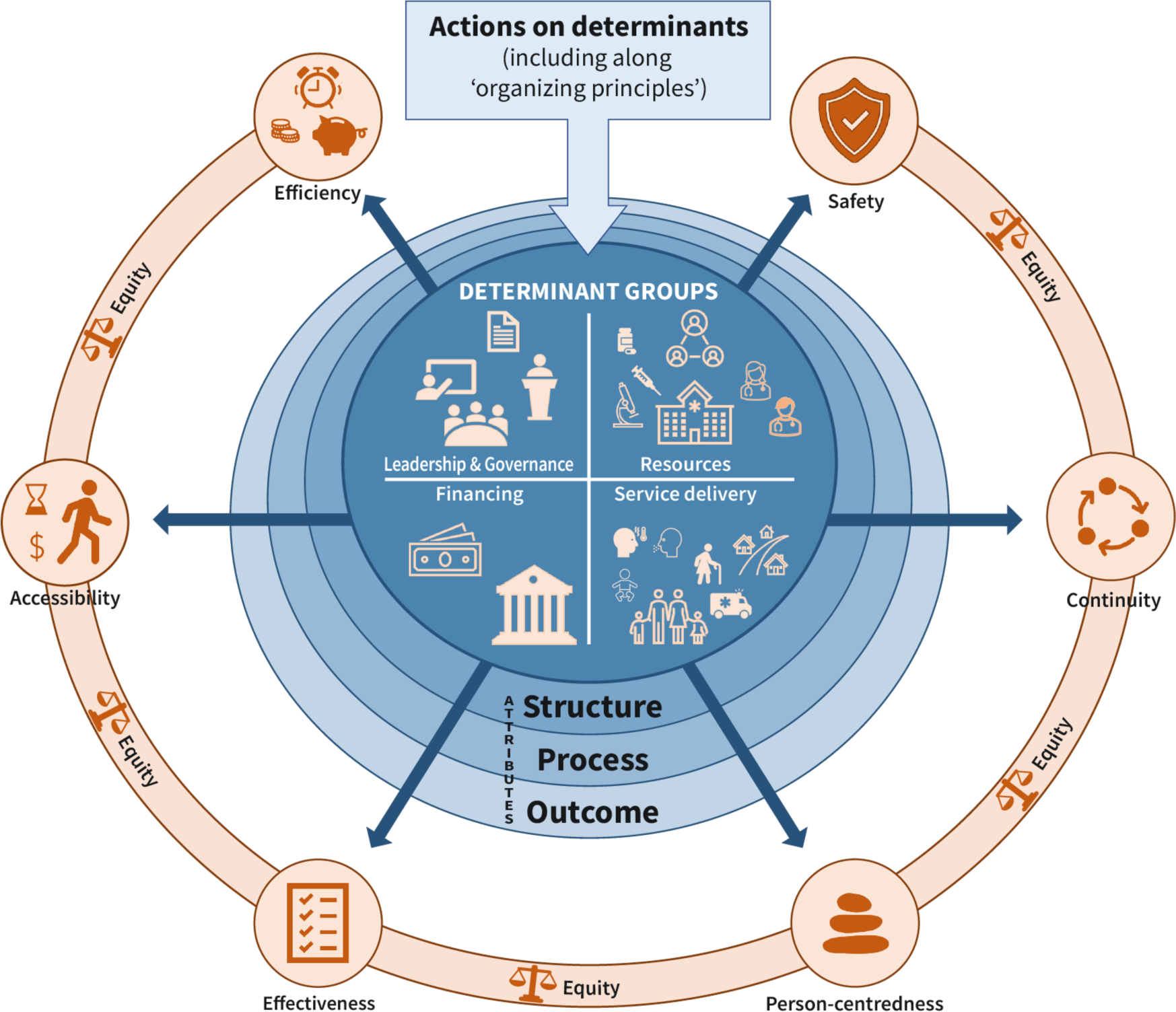
Framework for good quality care for chronic conditions

## DISCUSSION

Our results support creating a chronic care quality framework considering different individual and organizational perspectives, the whole gamut of chronicity from risks to complications. and the need for informal carers. These views are best expressed through our proposed determinant groups and specific actions that can be applied to specific steps in the healthcare journey of PwCC. The determinants and respective actions on certain determinants can contribute to the achievement of one or more of the seven chronic care quality aims. We have established that equity is an aim by itself as well as a cross-cutting one that should also be considered when achieving other aims. For instance, care that is effective should be made accessible regardless of personal characteristics of PwCC. Measuring achievement can be done by looking into specific indicators of the structures, processes, and outcomes of care.

We recognize that the determinants and actions listed from our scoping review and validated through the Delphi survey may not be exhaustive. However, our determinant groups are all-encompassing and include actors outside of the health system, as demonstrated in both our scoping review and Delphi results. Further to this, ‘journey consciousness’ reminds these actors that there is a collective responsibility for the best system performance along the PwCC journey and that everyone has to reflect how their contribution fits in the journey, keeping in mind that the state of the PwCC in front of them is determined by the history of the disease, and by how the health system has handled the PwCC and their risk-exposures and disease(s) in the past, is handling them right now, and how they will be handled in the future. Failure to provide good quality health services in an earlier step of the journey will redound on the succeeding steps. Journey consciousness and the (non-)achievement of good quality chronic care as related to each step can also be partly measured through cascades of care[85].

Starting the journey at the level of risk prevention brings to attention the actions that need to be done to prevent and control risks. Engagement and involvement of a wider range of actors is possible in the co-design and co-implementation of interventions on risks, and in addressing the social, structural and commercial determinants of the development and worsening of chronic conditions [34,39,40,44,60]. Such stakeholder-, community- and PwCC-engagements can be initiated at both the national and local levels. However, while actions on these ‘external’ determinants are expected to be made – or at the least, initiated – within the health system, there should be clear understanding of the responsibilities of the health system. Addressing the various risks and the social/structural/commercial determinants themselves, e.g. air pollution control, the regulation of sales of unhealthy products, food formulations, tobacco and alcohol taxations, etc. would need actions beyond the scope of the health system.

We also stress the importance of the PwCC (and their families and, where applicable, their informal caregivers), whom we identified as a determinant (a resource), in achieving good quality chronic care that starts from their own individual level. About 95-99% of care is given by the PwCC (or their families/ informal caregivers) to their own self; they are in-charge of their own health on a day-to-day basis [85]. Equipping PwCCs and their direct caregivers with self-management education and providing them support to self-manage will contribute to better chronic care. PwCCs can also be tapped as peer educators to provide self-management education and support, augmenting much-needed workforce.

Regarding monitoring of quality, achieving the aims of chronic care quality through their determinants and the actions are, to a fair extent, measurable. As illustrated in our CCCQ framework, this allows assessing whether quality of care is in place or has been acted upon. To look into specific and directly measurable variables of importance for quality of chronic care, our results are congruent with Donabedian’s structures, processes and outcomes framework [2]. Structure measures refer to the attributes of the health service or the healthcare provider, i.e. the presence or adequacy or magnitude of specific determinants, such as availability of services, health care provider to patient ratio, presence of guidelines. Process measures entail attributes relative to what the health system/service or the healthcare provider does to maintain or improve health, for instance if care delivery follows standards of care, healthcare workers follow standard operating procedures, referral/counter-referral mechanisms are used. Last, outcome measures are attributes that reflect the impact of the health service or the health care provided to the PwCC, for example improved glycemia in people with diabetes, reduced mortality rate. These attributes can have set criteria and/or indicators, e.g.: presence/availability of specific chronic care services, number of healthcare providers trained in self-management education provision (structure); number of healthcare providers following/utilising clinical practice guidelines (process); a decrease in glycosylated haemoglobin by certain percentage points (outcome), etc. Considering structures and processes expands measures of quality care beyond (clinical) outcomes. We do not give specific indicators or criteria in the CCCQ framework; rather, we indicate that attributes can be measured though structures, processes and outcomes. The measurable attributes listed in Table 2 give an idea of what indicators and criteria can be used; implementers can take inspiration from these examples and formulate their own measures based on their context and the specific interventions they implement.

## LIMITATIONS

We presented concepts relative to quality of care, tailored to chronic conditions. We note that there is very minimal literature available from low resource settings; we addressed this by ensuring representativity of low- and middle-income country expertise and experience in our Delphi survey.

While we have specified chronic care quality aims, determinants, actions and attributes from a scoping review and validated these with a Delphi survey, these warrant implementation research especially to identify what specific determinants need to be acted upon, what organising principles are warranted in the context to orient specific actions, and to tailor the structure, process, and outcome attributes that will be used to measure achievement of the aims.

## CONCLUSIONS

We have moved from a generic understanding of quality of care to one tailored to chronic conditions, considering various views of individuals and organizations. We have determined the scope of attention, one which values a comprehensive offer of healthcare services, addresses risks and determinants, ensures biopsychosocial well-being of PwCC, and gives importance to measurable attributes relevant to the PwCC and their families, to the community, and to the health system. With this view, we formulated a chronic care quality framework that looks into different determinants of chronic care quality and actions that could achieve the aims of good quality chronic care.

## IMPLICATIONS FOR POLICY & FURTHER RESEARCH

The CCCQ framework developed in this study could be used to identify the leverage points to be targeted by interventions aimed at improving quality of chronic care or to monitor along the causal pathway the effectiveness of such interventions. For instance, as a further round of discussion in the Delphi survey performed for this study, the framework was used as the basis for discussing healthcare purchasing arrangements and their possible effectiveness in improving quality of chronic care in low- and middle-income countries. The Delphi survey results on purchasing arrangements will be presented separately. These are all components of the larger program of work implemented by WHO, which focuses on purchasing arrangements as an instrument to improve quality of health services for chronic conditions. It is expected that member nations will take inspiration from this program of work. Actors active in chronic care may also be inspired by our specifications, in designing good quality chronic care services or working on improvement strategies thereto.

Our outputs relative to the framing of chronic care quality are conceptual. Operationalization for systematic improvements in quality of chronic care can be a next step, among others.

## Data Availability

All data produced in the present study are available upon reasonable request to the authors

## Acknowledgments

We thank John de Maesschalck, Institute of Tropical Medicine, Antwerp, Belgium, and Rafael Nalupta and Shreyashi Paik (student interns) for their assistance with the screening of scientific literature in the scoping review. The online Delphi survey (administration) was facilitated by Lynette Dominguez, independent consultant and supported, as for the digital platform, by Martin Erpicum, Mesylab, Liege, Belgium. We are grateful to the participants of our Delphi panel for their invaluable contributions. The authors are themselves alone responsible for the views expressed in this article, which do not necessarily represent the views, decisions, or policies of the World Health Organization.

## Author Contributions

GK conceptualized the full work, based on a terms of reference published by WHO, with contributions from WVDP, MGAA, and DK. The scoping review protocol was prepared by GK, with contributions from WVDP, MGAA, DK, MR and BM. The scoping review was conducted by GK, WVDP, MGAA, and DK. The Delphi survey protocol was prepared by GK and BM, with contributions from WVDP, MGAA, DK, and MR. The Delphi survey questionnaire was prepared by BM and GK, with contributions from MR. Analysis of the Delphi survey was conducted by BM, GK and MR. This manuscript was drafted by GK, with contributions from WVDP, MGAA, DK, MR and BM. All authors read and approved the final version.

## Disclosure Statement

None of the authors have any financial competing interests regarding this research.

## Ethics and Consent

The Delphi survey protocol and an amendment thereto were reviewed and approved by the Institute of Tropical Medicine, Antwerp Institutional Review Board (protocol number 1627/22). Briefing sessions, scheduled in two moments to accommodate time differences, were conducted to orient prospective participants to the study (https://www.youtube.com/watch?v=A7FLpdIB0xs&t=1s). Full informed consent was obtained prior to participation.

## Funding Information

This study, as part of a programme of work “Purchasing instruments to strengthen quality health services for chronic illnesses”, was commissioned by the WHO Kobe Centre and the WHO Department of Health Financing and Economics. The Kobe Group, which includes Hyogo Prefecture, Kobe City, the Kobe Chamber of Commerce and Kobe Steel, in Japan, contributed financially to the development and production of the research report.

## Paper context

- **Main findings**: *The nature of chronic conditions, the journey through healthcare of people with chronic conditions and how different groups view care quality redound on ensuing distinct chronic care needs, warranting specifications for good quality chronic care*.
- **Added knowledge**: *The concepts proposed in our chronic care quality framework are tailored to the natural history of chronic conditions and considers the journey through healthcare of a person with chronic conditions as well as views of different stakeholders, to provide a comprehensive guide in designing/implementing/evaluating quality chronic care services throughout the continuum*.
- **Global health impact for policy and action**: *This work, developed to guide further work on designing purchasing instruments to improve quality of chronic care, particularly in low- and middle-income countries, may also be a source of inspiration for other interventions aiming at improving quality of chronic care*.

## Notes

### Competing Interest Statement

The authors have declared no competing interest.

### Clinical Protocols

https://www.itg.be/en/research/research-themes/quality-of-care-for-chronic-conditions

### Author Declarations

The Delphi survey protocol and an amendment thereto were reviewed and approved by the Institute of Tropical Medicine, Antwerp Institutional Review Board (protocol number 1627/22).

